# Pediatric Long COVID Subphenotypes: An EHR-based study from the RECOVER program

**DOI:** 10.1101/2024.09.17.24313742

**Authors:** Vitaly Lorman, L. Charles Bailey, Xing Song, Suchitra Rao, Mady Hornig, Levon Utidjian, Hanieh Razzaghi, Asuncion Mejias, John Erik Leikauf, Seuli Bose Brill, Andrea Allen, H Timothy Bunnell, Cara Reedy, Abu Saleh Mohammad Mosa, Benjamin D Horne, Carol Reynolds Geary, Cynthia H. Chuang, David A Williams, Dimitri A Christakis, Elizabeth A Chrischilles, Eneida A Mendonca, Lindsay G. Cowell, Lisa McCorkell, Mei Liu, Mollie R Cummins, Ravi Jhaveri, Saul Blecker, Christopher B. Forrest, the RECOVER Consortium

**Affiliations:** Applied Clinical Research Center, Children’s Hospital of Philadelphia, Philadelphia, PA, USA; Department of Biomedical Informatics, Biostatistics and Medical Epidemiology (BBME), University of Missouri School of Medicine, Columbia, MO, USA; Department of Pediatrics, University of Colorado School of Medicine and Children’s Hospital Colorado, Aurora, CO, USA; RECOVER Patient, Caregiver, or Community Advocate Representative, New York, NY, USA; Department of Infectious Diseases, St. Jude Children’s Research Hospital, Memphis, TN, USA; Department of Psychiatry and Behavioral Sciences, Division of Child and Adolescent Psychiatry and Child Development, Stanford University School of Medicine, Palo Alto, CA, USA; Department of Internal Medicine College of Medicine, Division of General Internal Medicine, The Ohio State University, Columbus, OH, USA; Biomedical Research Informatics Center, Nemours Children’s Health, Wilmington, DE, USA; Department of Biomedical Informatics, Biostatistics, and Medical Epidemiology, University of Missouri School of Medicine, Columbia, MO, USA; Intermountain Medical Center Heart Institute, Salt Lake City, UT, USA; Department of Pathology, Microbiology, and Immunology, University of Nebraska Medical Center, Omaha, NE, USA; Penn State College of Medicine, Hershey, PA, USA; Department of Anesthesiology, University of Michigan, Ann Arbor, MI, USA; Center for Child Health, Behavior and Development, Seattle Children’s Research Institute, Seattle, WA, USA; Department of Epidemiology, College of Public Health, The University of Iowa, Iowa City, IA, USA; Division of Biomedical Informatics, Cincinnati Children’s Hospital Medical Center, Cincinnati, OH, USA; O’Donnell School of Public Health, The University of Texas Southwestern Medical Center, Dallas, TX, USA; Department of Health Outcomes and Biomedical Informatics, College of Medicine University of Florida, Gainesville, FL, USA; College of Nursing, University of Utah, Salt Lake City, UT, USA; Division of Infectious Diseases, Ann & Robert H. Lurie Children’s Hospital of Chicago, Chicago, IL, USA; Department of Population Health, New York University Grossman School of Medicine, New York, NY, USA

## Abstract

Pediatric Long COVID has been associated with a wide variety of symptoms, conditions, and organ systems, but distinct clinical presentations, or subphenotypes, are still being elucidated. In this exploratory analysis, we identified a cohort of pediatric (age <21) patients with evidence of Long COVID and no pre-existing complex chronic conditions using electronic health record data from 38 institutions and used an unsupervised machine learning-based approach to identify subphenotypes. Our method, an extension of the Phe2Vec algorithm, uses tens of thousands of clinical concepts from multiple domains to represent patients’ clinical histories to then identify groups of patients with similar presentations. The results indicate that cardiorespiratory presentations are most common (present in 54% of patients) followed by subphenotypes marked (in decreasing order of frequency) by musculoskeletal pain, neuropsychiatric conditions, gastrointestinal symptoms, headache, and fatigue.

## INTRODUCTION

Long COVID [or the closely related post-acute sequelae of COVID-19 (PASC)] is a condition characterized by persistence or development of symptoms or health conditions after SARS-CoV-2 infection; the initial CDC definition set a threshold of 4 or more weeks from acute infection[1]. Incidence estimates among pediatric patients who have had COVID vary substantially, depending on factors such as breadth of symptoms considered and how long they persist[2,3]. Studies of the clinical manifestations and underlying mechanisms of Long COVID point to a wide variety of symptoms, conditions, and body systems affected [2–10], and understanding of the specific subtypes is still developing.

Presentations of Long COVID may differ by both disease-specific and patient-specific factors, and accounting for these differences may be important for both Long COVID research and patient care. Long COVID studies in adult populations may not apply to children due to several factors, including symptom expression and attribution, marked age-related differences in the biology of the immune system, patterns of healthcare use, burden of comorbidities, and altered impact of social determinants of health. At the variable level, symptoms and conditions affecting the respiratory, circulatory, nervous, musculoskeletal, and digestive symptoms have been shown to occur significantly more frequently in the post-acute period following SARS-CoV-2 infection as compared with SARS-CoV-2 negative control cohorts. While this heterogeneity has been well-documented, less is known about patient-level co-occurrences of these symptoms and conditions in pediatric populations. Such an analysis would point to clinical subphenotypes of Long COVID, help clarify a more specific definition of Long COVID, and could provide insight into pathophysiological mechanisms and possible treatment responses that may be specific to certain clinical presentations of Long COVID.

Electronic Health Records (EHRs) provide a useful source of data for identifying Long COVID subphenotypes as they capture clinically relevant information for a large and longitudinal cohort of patients. Furthermore, the heterogeneity of Long COVID signs, symptoms, and health-related conditions suggests that subphenotypes may need to be identified by incorporating many potentially relevant variables, including diagnoses, procedures, and medications. EHR-based studies have identified Long COVID subphenotypes in adult populations[11,12]. Subphenotypes have also been characterized in children with Multisystem Inflammatory Syndrome in Children (MIS-C), a form of Long COVID by definition that is considered a distinct entity [13][14].

The goal of this study is to identify subphenotypes in a large cohort of pediatric patients with evidence of non-MIS-C Long COVID [15]. Prompted by the need to analyze a wide range of clinical variables to detect the many potential manifestations of Long COVID as well as their co-occurrences, we employ an unsupervised machine-learning method based on clinical concept embeddings, an extension of the Phe2Vec automated disease phenotyping algorithm, which is an adaptation of a natural language processing method to clinical data[16]. The foundation of our method is a concept embedding model trained from the clinical facts of 9.8 million patients to produce high-dimensional numeric representations of over 70 thousand unique diagnosis, procedure, and medication concepts. We then apply this model to represent and cluster the clinical trajectories of a cohort of pediatric patients with evidence of Long COVID.

## RESULTS

### Concept embedding model

For the first stage of our unsupervised machine learning-based pipeline for identifying Long COVID subphenotypes, we trained a concept embedding model from the clinical histories of 9,168,152 patients in the pediatric RECOVER EHR data source (Figure 1). This resulted in 200-dimensional vector representations of 77,337 concepts across the combined vocabularies of diagnosis, procedure, and medication codes.

**Figure 1:**
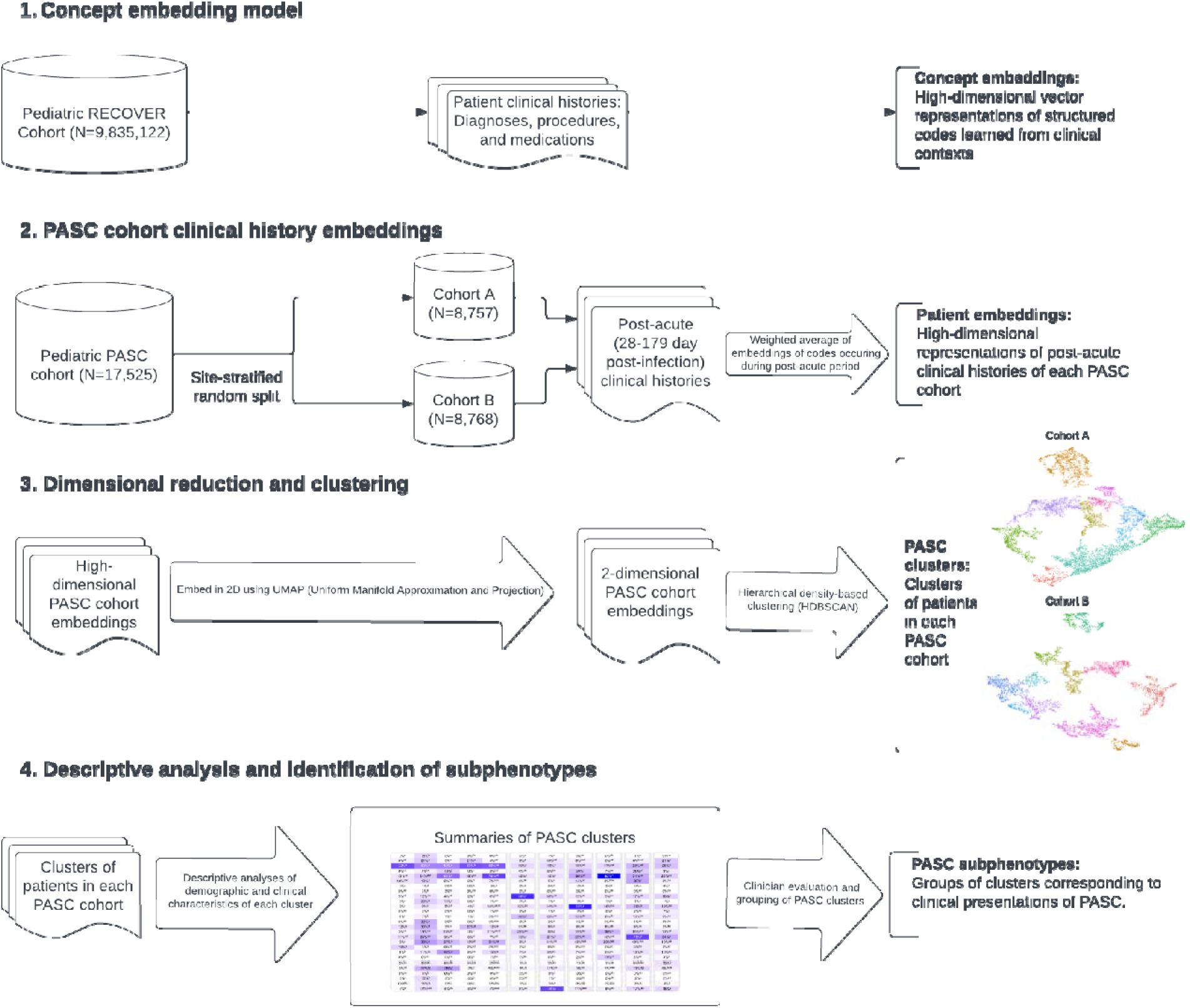
Subphenotype model flowchart.

### Long COVID cohort

There were 17,525 children and adolescents at 38 medical institutions identified as having evidence of Long COVID by a rules-based algorithm on the basis of a Long COVID diagnosis or post-acute evidence of Long COVID-associated diagnoses following evidence of SARS-CoV-2 infection [15]; patients with evidence of complex chronic disease prior to infection were excluded from the cohort. We refer to the cohort of patients with evidence of Long COVID as identified by this algorithm as the ‘Long COVID cohort.’ We randomly split this group into two cohorts (stratifying by site to ensure similar proportions of patients in each group), resulting in 8,757 in cohort A (training cohort) and 8,768 in cohort B (validation cohort), with similar distributions of all descriptive variables (Table 1). A plurality of patients in the ong COVID cohort were in the age 16-20 group (30.4% overall) and a majority were female (54.5%). Patients in this cohort were more likely to have been infected with SARS-CoV-2 during the November 2021-February 2022 period, coinciding with the Omicron wave, than in other time periods. Moderate and severe acute COVID-19 presentations were uncommon (4.9% and 3.2%, respectively). Thirty-seven percent of the cohort had evidence of at least one chronic disease that did not meet the definition of a complex chronic condition.

**Table 1:** Characteristics of Study Sample.

|  |  | **Overall**<br>**N=17,525** | **Cohort A**<br>**N=8,757** | **Cohort B**<br>**N=8,768** | **SMD** |
| --- | --- | --- | --- | --- | --- |
| **Age group (n/%)** | <1 | 1716 (9.8) | 873 (10.0) | 843 (9.6) | 0.028 |
|  | 1-4 | 2655 (15.1) | 1346 (15.4) | 1309 (14.9) |  |
|  | 5-11 | 3823 (21.8) | 1900 (21.7) | 1923 (22.0) |  |
|  | 12-15 | 4010 (22.9) | 1963 (22.4) | 2047 (23.4) |  |
|  | 16-20 | 5321 (30.4) | 2686 (30.6) | 2635 (30.1) |  |
| **Sex (n/%)** | Female | 9555 (54.5) | 4777 (54.5) | 4778 (54.6) | 0.002 |
|  | Male/Other/Unknown | 7970 (45.5) | 3991 (45.5) | 3979 (45.4) |  |
| **Race/ethnicity (n/%)** | Hispanic | 3956 (22.6) | 2005 (22.9) | 1951 (22.3) |  |
|  | Asian/PI | 570 (3.3) | 314 (3.6) | 256 (2.9) | 0.048 |
|  | Black/AA | 2394 (13.7) | 1164 (13.3) | 1230 (14.0) |  |
|  | Multiple | 388 (2.2) | 183 (2.1) | 205 (2.3) |  |
|  | Other/Unknown | 1481 (8.5) | 730 (8.3) | 751 (8.6) |  |
|  | White | 8736 (49.8) | 4372 (49.9) | 4364 (49.8) |  |
| **Cohort entry period (n/%)** | March-June 2020 | 356 (2.0) | 167 (1.9) | 189 (2.2) | 0.056 |
|  | July-October 2020 | 928 (5.3) | 492 (5.6) | 436 (5.0) |  |
|  | November-February 2021 | 2289 (13.1) | 1092 (12.5) | 1197 (13.7) |  |
|  | March-June 2021 | 1261 (7.2) | 622 (7.1) | 639 (7.3) |  |
|  | July-October 2021 | 2851 (16.3) | 1477 (16.8) | 1374 (15.7) |  |
|  | November-February 2022 | 6195 (35.3) | 3111 (35.5) | 3084 (35.2) |  |
|  | March-June 2022 | 2165 (12.4) | 1076 (12.3) | 1089 (12.4) |  |
|  | July-August 2022 | 1480 (8.4) | 731 (8.3) | 749 (8.6) |  |
| **ICU (acute) (n/%)** | 0 | 17220 (98.3) | 8621 (98.3) | 8599 (98.2) | 0.010 |
|  | 1 | 305 (1.7) | 147 (1.7) | 158 (1.8) |  |
| **Hospitalized (acute) (n/%)** | 0 | 16346 (93.3) | 8169 (93.2) | 8177 (93.4) | 0.008 |
|  | 1 | 1179 (6.7) | 599 (6.8) | 580 (6.6) |  |
| **COVID-19 acute phase severity of illness (n/%)** | Asymptomatic | 11093 (63.3) | 5524 (63.0) | 5569 (63.6) | 0.035 |
|  | Mild | 5004 (28.6) | 2540 (29.0) | 2464 (28.1) |  |
|  | Moderate | 865 (4.9) | 444 (5.1) | 421 (4.8) |  |
|  | Severe | 563 (3.2) | 260 (3.0) | 303 (3.5) |  |
| **Presence of existing chronic condition (n/%)** | 0 | 11034 (63.0) | 5542 (63.2) | 5492 (62.7) | 0.010 |
|  | 1 | 6491 (37.0) | 3226 (36.8) | 3265 (37.3) |  |

### Subphenotype Identification Pipeline

The subphenotype identification pipeline (consisting of applying the concept embedding model to the post-acute clinical histories of the Long COVID cohort) was first applied to cohort A to tune model hyperparameters; this is described in more detail in the supplement. The final model was then run once more on cohort A and on cohort B (the validation cohort). This resulted in 11 clusters identified in cohort A and 12 clusters in cohort B. Descriptive analyses, further described below, were performed on these clusters and are reported in the supplement (Figures 2S A-B and Tables 1S A-B). From these analyses, clusters were assigned clinically meaningful names and further grouped into subphenotypes. Cluster groupings which constitute the subphenotypes are shown in Tables 1S A-B and Figures 2S A-B. Finally, we performed descriptive analyses on the subphenotypes—for cohort B, these are shown in Table 2 and Figures 2 and 3.

**Figure 2:**
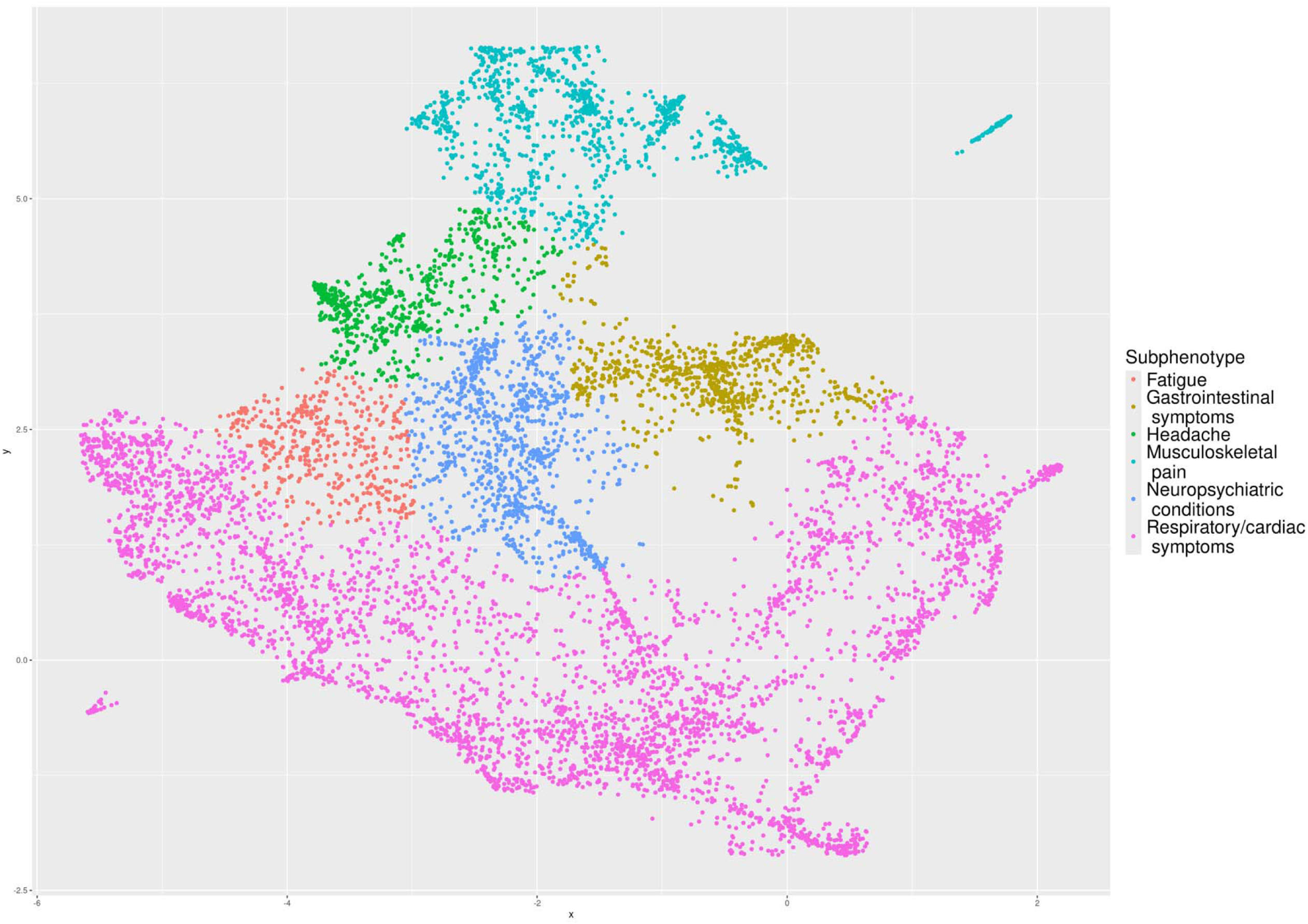
Subphenotype embeddings of PASC cohort B clinical histories.

**Figure 3:**
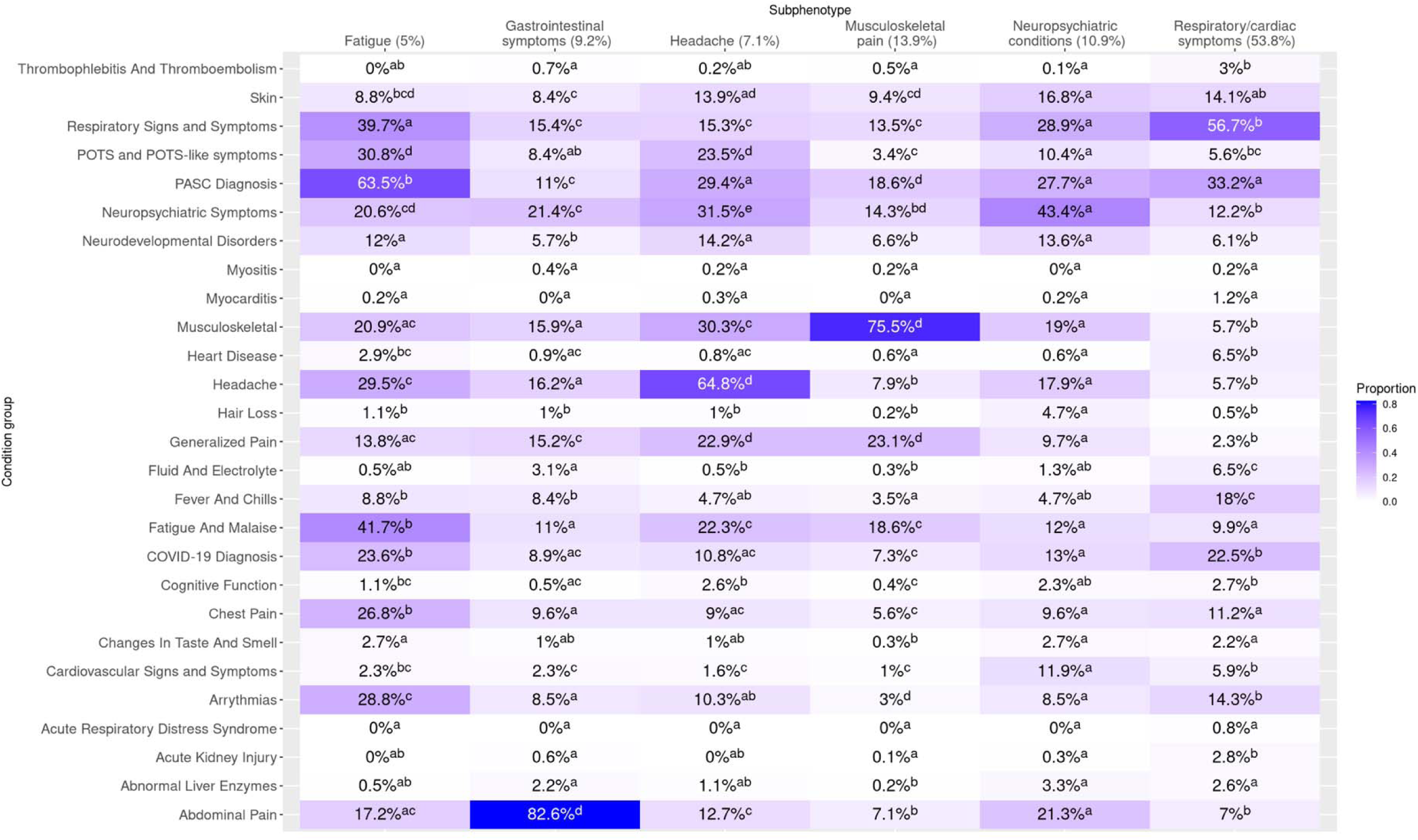
Heatmap of incident post-acute diagnoses, cohort B. **Legend:** Summary of clusters by presence of incident PASC-associated diagnoses. To be counted, diagnoses in the respective clusters had to occur in the 28-179 post-acute period following infection and not have been present in the 18 months prior. Cells display proportions of patients in the cluster with the corresponding PASC-associated diagnosis group, and the results of Compact Letter Display (CLD) analysis are represented in superscripts. For a given incident PASC-associated diagnosis group (row), two clusters share the same letter when proportions did not differ significantly (via multiple-testing adjusted chi squared testing) between the two clusters.

**Table 2:** Demographic and Clinical Characteristics of Subphenotype Groups in Cohort B. Note: cells marked with an asterisk have been modified by a random count between 0 and 4 to prevent reidentification of that cell or a cell in the same groups.

|  |  | Subphenotype |  |  |  |  |  |
| --- | --- | --- | --- | --- | --- | --- | --- |
|  |  | Fatigue<br>(441, 5.0%) | Gastrointestinal<br>symptoms<br>(810, 9.2%) | Headache<br>(620, 7.1%) | Musculoskeletal<br>pain<br>(1218, 13.9%) | Neuropsychiatric<br>conditions (952,<br>10.9%) | Respiratory/cardiac<br>symptoms<br>(4716, 53.85%) |
| **Age group**<br>**(n/%)** | <1 | 0 (0%) | 6 (0.7%) | 0 (0%) | 17 (1.4%) | 2 (0.2%)* | 816 (17.3%) |
|  | 1-4 | 2 (0.4%)* | 49 (6%) | 16 (2.6%) | 60 (4.9%) | 23 (2.4%)* | 1162 (24.6%) |
|  | 5-11 | 90 (20.4%)* | 233 (28.8%) | 116 (18.7%) | 315 (25.9%) | 190 (20%) | 978 (20.7%) |
|  | 12-15 | 145 (32.9%) | 206 (25.4%) | 233 (37.6%) | 419 (34.4%) | 298 (31.3%) | 746 (15.8%) |
|  | 16-20 | 204 (46.3%) | 316 (39%) | 255 (41.1%) | 407 (33.4%) | 439 (46.1%) | 1014 (21.5%) |
| **Sex (n/%)** | Female | 285 (64.6%) | 536 (66.2%) | 421 (67.9%) | 610 (50.1%) | 643 (67.5%) | 2283 (48.4%) |
|  | Male/Other<br>/Unknown | 156 (35.4%) | 274 (33.8%) | 199 (32.1%) | 608 (49.9%) | 309 (32.5%) | 2433 (51.6%) |
| **Race/ethnicity (n/%)** | Black/AA | 43 (9.8%) | 105 (13%) | 73 (11.8%) | 177 (14.5%) | 139 (14.6%) | 693 (14.7%) |
|  | Asian/PI | 11 (2.5%) | 19 (2.3%) | 6 (1%) | 22 (1.8%) | 29 (3%) | 169 (3.6%) |
|  | Hispanic | 69 (15.6%) | 183 (22.6%) | 100 (16.1%) | 193 (15.8%) | 251 (26.4%) | 1155 (24.5%) |
|  | White | 264 (59.9%) | 426 (52.6%) | 368 (59.4%) | 710 (58.3%) | 457 (48%) | 2139 (45.4%) |
|  | Multiple | 9 (2%) | 18 (2.2%) | 16 (2.6%) | 34 (2.8%) | 15 (1.6%) | 113 (2.4%) |
|  | Other/Unknown | 45 (10.2%) | 59 (7.3%) | 57 (9.2%) | 82 (6.7%) | 61 (6.4%) | 447 (9.5%) |
| **Cohort entry**<br>**period (n/%)** | Mar-Jun<br>2020 | 4 (0.9%)* | 21 (2.6%) | 10 (1.6%) | 44 (3.6%) | 19 (2%) | 92 (2%) |
|  | Jul-Oct 2020 | 17 (3.9%)* | 50 (6.2%) | 38 (6.1%) | 108 (8.9%) | 52 (5.5%) | 170 (3.6%) |
|  | Nov-Feb<br>2021 | 47 (10.7%) | 148 (18.3%) | 102 (16.5%) | 239 (19.6%) | 153 (16.1%) | 508 (10.8%) |
|  | Mar-Jun<br>2021 | 33 (7.5%) | 50 (6.2%) | 43 (6.9%) | 114 (9.4%) | 62 (6.5%) | 337 (7.1%) |
|  | Jul-Oct 2021 | 79 (17.9%) | 113 (14%) | 102 (16.5%) | 172 (14.1%) | 147 (15.4%) | 761 (16.1%) |
|  | Nov-Feb 2022 | 155 (35.1%) | 295 (36.4%) | 220 (35.5%) | 390 (32%) | 346 (36.3%) | 1678 (35.6%) |
|  | Mar-Jun 2022 | 59 (13.4%) | 79 (9.8%) | 59 (9.5%) | 89 (7.3%) | 112 (11.8%) | 691 (14.7%) |
|  | Jul-Aug 2022 | 47 (10.7%) | 54 (6.7%) | 46 (7.4%) | 62 (5.1%) | 61 (6.4%) | 479 (10.2%) |
| **ICU (acute) (n/%)** |  | 0 (0%) | 2 (0.2%)* | 5 (0.8%) | 7 (0.6%) | 6 (0.6%) | 139 (2.9%) |
| **Hospitalization (acute) (n/%)** |  | 4 (0.9%)* | 56 (6.9%) | 21 (3.4%) | 40 (3.3%) | 34 (3.6%) | 425 (9%) |
| **COVID acute phase severity of illness (n/%)** | Asymptomatic | 336 (76.2%) | 451 (55.7%) | 404 (65.2%) | 760 (62.4%) | 624 (65.5%) | 2994 (63.5%) |
|  | Mild | 89 (20.2%) | 287 (35.4%) | 184 (29.7%) | 392 (32.2%) | 290 (30.5%) | 1222 (25.9%) |
|  | Moderate | 11 (2.5%) | 62 (7.7%) | 23 (3.7%) | 49 (4%) | 28 (2.9%) | 248 (5.3%) |
|  | Severe | 5 (1.1%) | 10 (1.2%) | 9 (1.5%) | 17 (1.4%) | 10 (1.1%) | 252 (5.3%) |
| **Presence of existing chronic condition (n/%)** |  | 160 (36.3%) | 373 (46%) | 250 (40.3%) | 458 (37.6%) | 428 (45%) | 1596 (33.8%) |
| **Most common diagnoses** |  | U09.9: Post COVID-19 condition, unspecified (57.6%)<br>R53.83: Other fatigue (30.8%)<br>U07.1: Emergency use of U07.1 \| | R10.9: Unspecified abdominal pain (45.6%)<br>R10.84: Generalized abdominal pain (28.8%)<br>R10.13: Epigastric pain (21.2%) | R51.9: Headache, unspecified (37.9%)<br>U09.9: Post COVID-19 condition, unspecified (25.5%)<br>G89.29: Other chronic pain (18.1%) | G89.29: Other chronic pain (19.4%)<br>M62.81: Muscle weakness (generalized) (14.1%)<br>M25.561: Pain in right knee (14.0%)<br>B94.8: Sequelae of other specified | U09.9: Post COVID-19 condition, unspecified (24.0%)<br>F41.9: Anxiety disorder, unspecified (13.5%)<br>U07.1: Emergency use | U09.9: Post COVID-19 condition, unspecified (28.3%)<br>U07.1: Emergency use of U07.1 \| COVID-19 (22.3%)<br>R05.9: Cough, unspecified (18.8%)<br>R50.9: Fever, unspecified (16.3%)<br>R09.81: Nasal |
|  |  | COVID-19 (23.4%)<br>R51.9: Headache, unspecified (23.1%)<br>R42: Dizziness and giddiness (22.2%) | K59.00: Constipation, unspecified (21.2%)<br>R11.0: Nausea (19.1%) | R42: Dizziness and giddiness (16.3%)<br>F41.9: Anxiety disorder, unspecified (14.2%) | infectious and parasitic diseases (13.1%)<br>M25.562: Pain in left knee (13.1%) | of U07.1 \| COVID-19 (13.0%)<br>R51.9: Headache, unspecified (12.2%)<br>R10.9: Unspecified abdominal pain (10.2%) | congestion (11.2%) |

### Cluster and subphenotype characterization

For each identified cluster (and subsequent subphenotype), we calculated the proportions of patients with diagnoses in 25 groups of Long COVID-associated condition groups (Figure 3, Figures 2S A-B). To differentiate presentations, we used Bonferroni-adjusted pairwise chi^2^ testing for each combination of condition group and patient group (cluster, subphenotype), presented in a compact letter display (CLD) format superimposed over the heatmaps. Additionally, we summarized patient characteristics and the most common diagnoses in each cluster and subphenotype (Table 2, Tables 1S A-B). Results for cohort B are described below—corresponding results for cohort A are shown in the supplement. We additionally summarized utilization patterns (Figure 6S) and presence of pre-existing (non-complex) chronic conditions (Figure 7S).

Cardiorespiratory presentations were most common, representing 53.8% of patients. This subphenotype (“Respiratory/cardiac symptoms”) was characterized by a statistically significantly greater proportion (56.7%) of respiratory diagnoses than in any other subphenotype (Figure 3). The respiratory/cardiac subphenotype is further stratified into six cluster-specific presentations differing by severity, age, post-acute utilization trajectories, and predominance of upper versus lower respiratory diagnoses (Table 1S-B and Figure 2S-B, Figure 6S).

Pain-related diagnoses distinguished a subphenotype (“Musculoskeletal pain”) representing 13.9% of patients, with “other chronic pain” as the most common diagnosis code. A third subphenotype (“Neuropsychiatric conditions”, representing 10.9% of patients) was characterized by a statistically significantly greater proportion of neuropsychiatric condition diagnoses than other subphenotypes, with anxiety disorder as the most common specific diagnosis. A fourth subphenotype (“Gastrointestinal symptoms”, representing 9.3% of patients) was characterized by a statistically significantly greater proportion of gastrointestinal diagnoses than other subphenotypes. A fifth subphenotype, “Headache” (representing 7.1% of patients), was characterized by a statistically significantly greater proportion of headache diagnoses than other subphenotypes—neuropsychiatric diagnoses were relatively more common in this subphenotype as well, and patients with this subphenotype had the highest volume of post-acute utilization with the exception of a more severe lower respiratory cluster (Figure 6S). Finally, a subphenotype (“Fatigue”, representing 5.0% of patients) was characterized by statistically significantly greater proportions of both fatigue and malaise diagnoses (41.7%) as well as Long COVID diagnoses (63.5%); diagnoses of chest pain, arrythmias, and respiratory signs and symptoms were common in this subphenotype as well.

### Comparison to cohort A and to a matched control cohort

Subphenotypes identified in cohort B resembled those in Cohort A (Figures 1S, 4S). Cohort A had five cardiorespiratory clusters constituting a respiratory/cardiac subphenotype representing 50.8% of patients. At the more granular level, the clusters in cohort A had similar characterizations (by severity, age, upper vs lower respiratory) as those in cohort B. Subphenotypes representing musculoskeletal pain, neuropsychiatric conditions, gastrointestinal symptoms, headache, and fatigue were identified in similar proportions in cohort A. The fatigue cluster in cohort A had a relatively smaller proportion of fatigue and malaise diagnoses than the corresponding cluster in cohort B and a great proportion of non-specific Long COVID diagnoses; cardiac diagnoses were also relatively less common in this cluster than in the corresponding fatigue cluster in cohort B.

A cohort of patients with no evidence of COVID-19 (with index date defined by a random visit during the study period) was matched to cohort B using nearest neighbor propensity score matching on sex, racial and ethnic group, time period of index date, institution, and presence of chronic condition across each of the PMCA body systems. Patients with a history of complex chronic conditions were similarly excluded from this control cohort. These covariates were well-balanced after matching (absolute SMDs all less than 0.1). There were 16 clusters identified in this control cohort; Figure 3S shows proportions of diagnoses in each of the 25 Long COVID-associated diagnosis groups. A visualization of cluster centroids as identified in cohorts A, B, as well as the matched control cohort is shown in Figure 4S and enables comparison of subphenotypes/clusters identified in the three cohorts.

### Demographic characteristics of clusters

There were substantial demographic differences in our reported subphenotypes (Table 2), most stark at the more granular cluster level (Table 1S-B). Patients younger than 4 years were primarily represented in the respiratory/cardiac subphenotype, particularly in the lower respiratory, more severe and upper respiratory clusters. Conversely, older children were primarily represented in the lower respiratory (0B, 6B), upper respiratory-inflammatory (7B), and non-respiratory clusters. Female patients were relatively underrepresented in the respiratory/cardiac and musculoskeletal pain subphenotypes and overrepresented in the fatigue, headache, gastrointestinal, and neuropsychiatric subphenotypes. Relative to the full cohort, Hispanic patients were overrepresented in the respiratory/cardiac subphenotype (particularly upper respiratory, inflammatory-younger and lower respiratory, more severe clusters) and the neuropsychiatric conditions subphenotype. Non-Hispanic Black/African-American patients were overrepresented in the neuropsychiatric conditions, musculoskeletal pain, and respiratory/cardiac subphenotypes (particularly the lower respiratory--more severe, upper respiratory--obstructive, and upper respiratory—inflammatory, younger clusters). Non-Hispanic Asian/Pacific-Islander patients were relatively overrepresented in the respiratory/cardiac subphenotype. Non-Hispanic White patients were overrepresented in the fatigue, headache, and musculoskeletal pain subphenotypes.

### Sensitivity analyses

The results of a sensitivity analysis in which patients who were not assigned a cluster were left unclustered (rather than assigned the cluster of the nearest centroid) are shown in Figure 5S.

## DISCUSSION

The heterogeneity of specific Long COVID features in pediatric populations has been catalogued in a number of prior studies [5–7,9,17,18]. In this study, we extended this work by characterizing patterns of symptoms, signs, medications, and procedures that suggest specific subphenotypes of Long COVID in children and adolescents who do not have evidence of existing complex chronic conditions. We applied and extended the Phe2Vec algorithm [16] to the problem of subphenotyping, leveraging the high-dimensional nature of EHR data.

Our model identified six subphenotypes, with cardiorespiratory presentations identified in about half of patients. Other common subphenotypes in order of frequency included musculoskeletal pain, neuropsychiatric conditions, gastrointestinal symptoms, headaches, and fatigue. Each subphenotype was well-differentiated by a specific group of diagnoses, suggesting that distinct populations may manifest these specific Long COVID presentations. Within the respiratory/cardiac subphenotype, we identified five more specific clusters, with presentations differentiated by both clinical (upper versus lower respiratory, severe and less severe, obstructive and inflammatory) as well as demographic characteristics (Table 1S-B). Interestingly, patients with more severe acute infection were classified almost entirely in the respiratory/cardiac subphenotype (specifically, within the lower respiratory, severe cluster).

The fatigue subphenotype was somewhat more heterogeneous; in addition to fatigue, cardiac diagnoses (chest pain and arrythmias), headaches, musculoskeletal pain, neuropsychiatric symptoms, and POTS-like symptoms such as dizziness and giddiness were relatively more common, as well as non-specific Long COVID diagnoses. Although these more common groups of diagnoses did not always occur in the same sets of patients, this constellation of diagnoses is suggestive of myalgic encephalomyelitis/chronic fatigue syndrome (ME/CFS)[19]. Although specific diagnostic codes for ME/CFS exist, and a new ICD-10-CM code was introduced on 1 October 2023, the disease remains very likely to be under-diagnosed, particularly in children [20–22]. In addition, because clinical criteria for ME/CFS require symptoms to persist for a minimum of 6 months from onset before assigning the diagnosis, the use of a 28-to-179-day observational window following the index infection in this study made it impossible to strictly meet the 6-month criterion for establishing an ME/CFS diagnosis[23]. It is possible that our fatigue subphenotype identifies patients with ME/CFS from the above non-specific diagnoses often associated with it. The heterogeneity of this subphenotype may also be responsible for the proximity of this subphenotype to cardiorespiratory presentations (Figure 4S) and the somewhat different characterization of this subphenotype in Cohort A (Figure 1S). Further work is necessary to analyze the clinical characterization of this subphenotype and its reproducibility.

While fatigue was the most commonly reported Long COVID feature in some studies[6,7], we found cardiorespiratory presentations to be the most common subphenotype, with the fatigue subphenotype above representing only about 5% of patients. However, diagnoses of fatigue were present across multiple subphenotypes (particularly the headache and musculoskeletal pain subphenotypes, in addition to the fatigue subphenotype). This suggests that fatigue often presents not in isolation but in combination with other aspects of Long COVID and may be present across multiple Long COVID manifestations. Other prospective studies may be able to capture fatigue more reliably than EHR data sources.

Commonly reported Long COVID manifestations not clearly identified in this study included a distinctly cardiac subphenotype and neurocognitive difficulties (commonly referred to as brain fog). In the case of a cardiac subphenotype, as patients with Multisystem Inflammatory Syndrome in Children (MIS-C) were excluded from analysis, this suggests that uniquely cardiac presentations may have substantial overlap with the MIS-C-affected population. Reported Long COVID manifestations such as neurocognitive difficulties/so-called “brain fog” lack a clear diagnosis and thus may be under-ascertained in EHR data, particularly among children. Finally, as our analysis only identified clusters present in at least 2% of the population, less common subphenotypes may have instead been grouped with others.

Compared to Long COVID subtypes identified in EHR-based studies in adult populations [11,12], we found both overlap and differences in Long COVID presentations. As in both studies, we found a more severe cardiorespiratory cluster within our respiratory/cardiac subphenotype with patterns of symptoms overlapping with the multisystem+lab cluster in reference [11] as well as the cardiac/renal subtype in reference [12]. Manifestations observed in specific subphenotypes in our study, particularly musculoskeletal, gastrointestinal, and neuropsychiatric symptoms, were more likely to be part of composite subphenotypes in studies in adult populations; for instance, the musculoskeletal pain subphenotype we observed is part of composite subphenotypes, grouped with fatigue in reference [11] and with headaches and sleep-wake disorders in reference [12]. This may be a result of different methods and particularly different levels of granularity in grouping similar presentations in the two studies but may also point to more specific Long COVID manifestations in pediatric populations. Similarly, the presence of multiple cardiorespiratory subtypes in this study may point to greater heterogeneity in respiratory manifestations of Long COVID in pediatrics.

Clustering methods have also been applied to PASC-probable patients in school-age (6-11 years) and adolescent (12-17 years) prospective cohorts [24]. Compared to these results, we also identified a cluster with high symptom burden (our lower respiratory-more severe cluster within the respiratory/cardiac subphenotype) and a predominantly gastrointestinal subphenotype. A headache and fatigue cluster in reference [24] resembles both our headache and fatigue subphenotypes. Differences include the predominance of a respiratory/cardiac subphenotype in our work which was not identified in reference [24]. We note that our respiratory/cardiac subphenotype was effectively the only one identified in age 0-4 patients, a population not included in [24]. Additionally, a subphenotype characterized by neuropsychiatric conditions was identified in our work but not in reference [24]. Conversely, a cluster characterized by loss of taste and smell was identified in the adolescent cohort and a cluster characterized by sleep impacts was identified in the school-age cohort in reference [24]. Differences between our findings may be due to several factors: difference in age groups select (age 0-20 in our study versus 6-17 years in [24]) and other cohort inclusion and enrollment criteria, granularity and definitions of variables used in clustering (individual diagnosis, medication, and procedure codes from EHR data versus presence of 89 symptoms collected by survey).

Our subphenotype classifications varied by age, sex, race, and ethnicity. Children younger than 4 were almost exclusively assigned to the respiratory/cardiac subphenotype (Table 3), mainly divided between a more severe lower respiratory cluster (characterized by greater frequency of arrythmias, fluid and electrolyte disturbances, hospitalizations, and ICU admissions) and a less severe upper respiratory cluster (characterized by cough, fever, and nasal congestion). This may indicate that primarily respiratory manifestation of Long COVID affect younger children, may reflect subsequent respiratory infections, or reflect general patterns of utilization in younger children, or may be a consequence of limitations in parental or child self-reporting of other kinds of symptoms (e.g., headaches or symptoms of anxiety disorders). Further, Hispanic and non-Hispanic non-White patients were overrepresented in the respiratory/cardiac subphenotype, a finding that has been corroborated in other studies [25]. Further exploration of these differences in presentation by sociodemographic characteristics is needed to determine whether patterns reflect differences in pathophysiology, symptom reporting, healthcare access, or utilization.

Although patients with complex chronic conditions (e.g., patients with actively treated cancer, muscular dystrophy, etc.) were excluded from this study due to the difficulties in attributing post-acute symptoms to COVID-19 versus existing conditions, patients with an existing non-complex chronic condition were overrepresented in the gastrointestinal and the neuropsychiatric conditions subphenotypes (Table 2); this may be suggestive of specific, as yet undetermined risk factors for these subphenotypes, or that these presentations manifest as exacerbations of existing chronic conditions evidenced by incident post-acute diagnoses.

Results from Cohort A (development cohort) were largely similar to those in Cohort B (validation cohort), with all six subphenotypes present in similar proportions in both cohorts (Figures 3, 1S), adding validity to our approach. At a more granular level, Cohort A exhibited a similar stratification into cardiorespiratory clusters, though one additional cluster characterized by a high proportion of non-specific Long COVID diagnoses (96.1%) was also identified. Other differences observed between the two cohorts were in the characterization of the fatigue subphenotype noted above, as well as the presence of two distinct neuropsychiatric conditions clusters in Cohort A versus one in cohort B, and the presence of two distinct musculoskeletal pain clusters in Cohort B versus one in cohort A. The heterogeneity of diagnosis, procedure, and medication codes associated with Long COVID together with relative overlap between different subphenotypes (e.g., presence of fatigue diagnoses across multiple subphenotypes) as well as potential difficulty in assigning subphenotypes to patients with less specific presentations may be responsible for these differences between the two cohorts.

The analysis of a matched control cohort with no evidence of SARS-CoV-2 infection produced 16 clusters representing a variety of clinical presentations. Distances between centroids of clusters in the control cohort compared to centroids of subphenotypes from the two Long COVID cohorts in Figure 4S show that our neuropsychiatric and gastrointestinal subphenotypes appear relatively near to clusters identified in the matched control cohort; conversely, the headache, fatigue, and respiratory/cardiac subphenotypes are relatively further from any clusters in the matched control cohort. This may indicate that headache, fatigue, and respiratory/cardiac subphenotypes of Long COVID are characterized by presentations that may be easier to differentiate from other clinical entities, whereas neuropsychiatric and gastrointestinal symptoms in patients with Long COVID may present more similarly to those exhibited in a more general care-seeking population, and therefore be more difficult to detect. This finding may also be a result of the limited ability of the diagnostic codes, prescriptions, and procedure codes that were used as input features to the Phe2vec model to describe any differences between COVID-associated and non-COVID associated neuropsychiatric or gastrointestinal disease.

Strengths of this study include use of large multi-site longitudinal EHR data; this enabled us to train a concept embedding model from a sufficiently large cohort so as to represent the semantic content of tens of thousands of concepts based on the clinical data of 9.1 million patients with greater generalizability than models trained on data from a single institution. Further, the novel concept-embedding-based methods for subphenotyping developed in our study allow us to effectively leverage the great variety of data available in EHRs by bringing it to bear on the study of pediatric Long COVID, a particularly heterogeneous condition. In place of alternative approaches in which variable definitions and groupings in the study of co-occurrence involve extensive curation of study variables which may be the source of study bias, concept similarity is learned from context in tens of millions of clinical encounters.

Our study has multiple limitations worth noting. First, the lack of a clinical case definition of Long COVID and corresponding ‘gold standard’ cohort meant we were reliant on the clinical rules-based phenotype developed in [15]. The Long COVID phenotype algorithm may under-identify patients because of low rates of use (i.e., mild cases) or physician underdiagnosis of symptoms, or it may produce false positives. The pattern of subphenotypes we identified, however, is consistent with the most commonly reported Long COVID symptoms in children [17,18], which lends plausibility to our findings. A second limitation, related to the first, is our use of the 28-179 day period following infection for identifying post-acute symptoms; symptoms of Long COVID can chance or first appear past the 6 month mark and may take longer to be captured in EHRs due to long waits to see a specialist. Our choice of a 6 month cutoff was motivated by the increased risk of misattributing symptoms that occur more than six months after the index infection as evidence of Long COVID; however, further research is necessary to understand how Long COVID presentations vary over time and how these are captured in EHRs. Second, EHR data reflects symptoms and conditions managed by clinicians, and if patients do not seek or have access to quality care, those data will be missing. A third limitation is the absence of patient laboratory testing results as an input to our pipeline; while results of laboratory testing may provide valuable information about patients’ Long COVID trajectories, early attempts to use these data mainly clustered patients by volume of utilization (grouping patients into those with high and low frequency of labs) and further investigation is necessary to make effective use of laboratory testing in concept embedding models. Fourth, as discussed above, subphenotypes which are less common (present in less than 2% of our cohort) or poorly captured in EHRs (e.g., “brain fog” or attentional problems, difficulties in school) are less likely to be detected. Augmenting structured data with physician notes (i.e., text) is a promising direction for capturing these symptoms and subtypes in the future. Fourth, the exclusion of patients with complex chronic disease from this study due to difficulties in attribution of symptoms means that subtypes of Long COVID defined by worsening of trajectories related to specific chronic conditions are less likely to be detected.

Methodologically, our concept embedding pipeline is an unsupervised algorithm; the lack of a gold standard dataset labeling patients with subtypes is a challenge for identifying the accuracy of our approach. While tuning pipeline hyperparameters on cohort A and reproducing clinically similar clusters in cohort B adds plausibility to our results, data from ongoing observation cohort studies has the potential to provide more accurate classification of Long COVID into subtypes and is a promising area for future work.

## ONLINE METHODS

### Data source

This retrospective cohort study is part of the NIH Researching COVID to Enhance Recovery (RECOVER) Initiative, which seeks to understand, treat, and prevent the post-acute sequelae of SARS-CoV-2 infection (22). The RECOVER EHR population includes clinical data for patients at 38 hospital systems across the United States. Data were extracted from version 11 of the pediatric RECOVER database, comprising 9,835,122 patients with evidence of testing or immunization for SARS-CoV-2 or diagnoses of COVID-19 or other respiratory illnesses between January 2019 and December 2022. Institutional Review Board (IRB) approval was obtained under Biomedical Research Alliance of New York (BRANY) protocol #21-08-508. As part of the BRANY IRB process, the protocol has been reviewed in accordance with institutional guidelines. BRANY waived the need for consent and HIPAA authorization.

### Study sample

Although there is a single ICD-10-CM code (U09.9) for post COVID-19 condition, unspecified (introduced 1 October 2021), it is not consistently applied in pediatrics. Consequently, use of the diagnosis code alone may not produce a representative cohort of patients with Long COVID. To define a larger and more representative cohort of patients with evidence of Long COVID, we used the PEDSnet rules-based computable phenotype for Long COVID[15]. The algorithm selects SARS-CoV-2 positive patients who had diagnoses during the 28-to-179-day post-acute period following infection of either direct clinician-diagnosed Long COVID (ICD-10-CM U09.9), or incident diagnoses associated with Long COVID in prior studies[5,17]. SARS-CoV-2 positive patients are identified by PCR, antigen and serology testing as well as the presence of COVID diagnosis codes and prescriptions of the COVID-specific medications nirmatrelvir/ritonavir and remdesivir. The index date of SARS-CoV-2 positivity is defined as the date of first positive test or COVID diagnosis. For patients who only had a diagnosis of Long COVID (U09.9) or Sequelae of other specified infectious and parasitic diseases (B94.8) with no prior SARS-CoV-2 test or COVID-19 diagnosis, the index date is imputed as a random date between 28 and 90 days prior to U09.9 or B94.8 diagnosis. Finally, due to the difficulties in attributing symptoms to Long COVID among patients with complex chronic conditions (as computed by Version 2.0 of the Pediatric Medical Complexity Algorithm (PMCA) [26]), patients with evidence of a complex chronic condition in the three years prior to cohort entry were excluded from the cohort. A flowchart describing the cohort definition is shown in Figure 2 of reference [15] and the full set of Long COVID-associated features used in the phenotype is listed in the supplementary appendix. Clinical histories of patients in this cohort were studied during the 28-to-179-day period following the SARS-CoV-2 positivity index date; we use ‘post-acute period’ to refer to this time period relative to infection throughout the manuscript.

A matched control cohort of patients with no evidence of COVID was identified and is further described in the supplement.

### Long COVID subphenotype pipeline

The ased pipeline used to identify Long COVID subphenotypes in our cohort is outlined in Figure 1. Below, we give brief descriptions of the main steps; a more technical description, including hyperparameters and methods for validation, is included in the supplement. The first two steps follow the approach of the Phe2Vec algorithm for EHR-based automated phenotyping.

#### Concept embedding model

Due to the large number of potentially relevant variables across the condition, drug, and procedure domains, we began by constructing numerical (vector) representations of the relevant clinical concepts. In the field of natural language processing, word embedding models are often used to produce such representations of words in such a way that the sematic relationships between words are encoded in their vector representations (e.g., words with similar meanings are represented by vectors that are close together). The Phe2Vec algorithm adapts these models, particularly the Word2Vec algorithm [27] to structured clinical data. In this analogy, words correspond to clinical concepts (represented by domain-specific structured codes, e.g., ICD10CM codes for diagnoses) and sentences correspond to concatenations of clinical concepts that are recorded in patients’ clinical histories over a given time period.

We trained the concept embedding model from the clinical histories of 9,168,152 patients in the pediatric RECOVER EHR data source. Our model uses codes from the following domains and vocabularies: conditions (ICD10CM), drugs (RxNorm Clinical Drug Forms), and procedures (ICD10PCS, HCPCS, CPT4). We constructed for each patient and each month-long period of their clinical history, a sentence consisting of the codes which occurred for that patient during that time period arranged in randomly permuted order. This resulted in a corpus of 99,413,139 sentences and yielded vector representations for a combined vocabulary of 77,337 concepts.

#### Patient clinical history embeddings

Equipped with the vectors representing structured clinical concepts, the next step in our approach was to extend this model to produce similar representations for the post-acute clinical histories of our Long COVID cohort. To construct these, we first identified the codes in our vocabularies that occurred in this cohort during the 28 to 179 day period following the index date. We restricted attention to only those codes that did not occur previously in the 7 day to 6 month washout period prior to the index date. We then assembled these codes for each patient in random order into sentences of codes. To construct vector representations of these sentences from the already-learned vector representations of codes (corresponding to words) we used the Simple but Tough-to-Beat Baseline for Sentence Embeddings [28]. The resulting 200-dimensional vectors represent the post-acute clinical histories of our cohort.

At this point, our approach deviated from that of Phe2Vec; while the goal of Phe2Vec is to phenotype patients by computing similarity between patient clinical trajectories and a given set of seed codes, our next step consisted of clustering the Long COVID cohort.

#### Dimensional reduction and clustering

While 200-dimensional space is more appropriate for embedding the full set of codes in our vocabulary, we found that the sparsity of representations of the post-acute histories of our smaller cohort of Long COVID patients impeded effective clustering (‘the curse of dimensionality’). As a result, prior to clustering we applied the UMAP algorithm[29] to embed the 200-dimensional vector representations of our cohort into 2-dimensional space; we additionally chose 2-dimensional space to facilitate visualization of the embedded representations. The irregular shapes and varying densities of clusters produced by the UMAP algorithm were not well-suited for k-means or hierarchical clustering algorithms; as such, we elected to use the density-based clustering algorithm HDBSCAN[30] to identify clusters in the embedded vector representations of the post-acute clinical histories of our cohort after their index infection. Because HDBSCAN does not always assign a cluster and allows some data to remain unclustered, we assigned unclustered patients to the cluster whose centroid was nearest to them. Thus, in our main analysis, each patient was assigned to (exactly) one cluster. We also conducted a sensitivity analysis in which we allowed patients to remain unclustered.

#### Hyperparameters, model selection, and validation

For model validation, a 50/50 random split into cohorts A and B was used; to better control for site-heterogeneity in code usage, we used a site-stratified split (i.e., resulting in equal distributions of patients across sites in the two groups). Pipeline hyperparameters were selected by running the pipeline and by comparing output on Cohort A (described further in the supplementary appendix). The final pipeline was then run on both cohorts.

### Descriptive and statistical analyses

To summarize the resulting clusters of patients in our cohort, we calculated, for each of 25 groups of Long COVID-associated conditions (each defined by a collection of diagnosis codes chosen by investigators based on prior work [5,15,17]), the proportions of patients in each patient cluster with an incident (using the same washout period above) diagnosis of that feature during the post-acute period. We represented these proportions using heatmaps, limiting them to groups of conditions which were represented in at least 20% of patients from at least one cluster. To differentiate presentations represented by patient clusters, we used Bonferroni-adjusted pairwise χ^2^ testing for each combination of condition group and patient clusters, presented in a compact letter display (CLD) format superimposed over the heatmap. In this presentation, for each Long COVID-associated feature (corresponding to a row in the heatmap), two patient clusters (corresponding to columns) share a letter in common exactly when proportions of patients with that feature did not significantly differ between the two patient clusters.

We further summarized the patient clusters by patient characteristics including age, sex, and race/ethnicity. Additionally, we used the Pediatric Medical Complexity Algorithm to compute presence of chronic conditions in the 3 years prior to index date; as complex chronic patients were excluded, only non-complex chronic patients (e.g., those with non-progressive and non-malignant conditions affecting only one body system, e.g., asthma) were summarized. We computed the proportion of patients with presence of chronic condition across 17 body systems. We also employed the acute pediatric COVID-19 severity typology developed in reference [31] to categorize patients’ acute infections as asymptomatic, mild (presence of symptoms), moderate (moderately severe COVID-19-related conditions such as gastroenteritis and pneumonia), and severe (unstable COVID-19-related conditions, ICU admissions, or mechanical ventilation); we then summarized proportions of patients by severity of infection in each cluster. We also summarized clinical trajectories by subphenotype over time by three utilization-based metrics: number of distinct visits per month, number of distinct providers seen per month, and number of body systems affected by month. Finally, we used the descriptive analyses above—particularly the distinguishing groups of symptoms and conditions and most common individual diagnoses in each cluster—to assign clinically descriptive names to each of the cluster; additionally, we grouped clinically similar presentations represented by the patient clusters into Long COVID subphenotypes.

### Sensitivity analyses

To assess the effect of allowing some patients to remain unclustered, we conducted an additional sensitivity analysis in which patients not assigned a cluster by HDBSCAN were left unclustered; we reproduced the descriptive and statistical analyses above for this assignment of clusters.

### Code and availability

Analyses were conducted using R version 4.0 and Python version 3.8.16. We used the following Python libraries: Gensim [32] for training the concept embedding model using the Word2Vec algorithm, UMAP [29] for dimensional reduction, and HDBSCAN[30] for clustering. Sentence embeddings were computed using Python code accompanying A Simple but Tough-to-Beat Baseline for Sentence Embeddings [28], and propensity score matching was conducted using the R MatchIt package [33]. Code used to implement the subphenotype pipeline and produce the results of this manuscript is available at https://github.com/PEDSnet/recover_pasc_subphenotype_manuscript.

## Disclosures

### Disclaimer

This content is solely the responsibility of the authors and does not necessarily represent the official views of the RECOVER Initiative, the NIH, or other funders.

## Data Availability

The results reported here are based on detailed individual-level patient data compiled as part of the RECOVER Program. Due to the high risk of reidentification based on the number of unique patterns in the date, patient privacy regulations prohibit us from releasing the data publicly. The data are maintained in a secure enclave, with access managed by the program coordinating center to remain compliant with regulatory and program requirements. Please direct requests to access the data, either for reproduction of the work reported here or for other purposes, to.

https://github.com/PEDSnet/recover_pasc_subphenotype_manuscript

## Acknowledgements

This study is part of the NIH Researching COVID to Enhance Recovery (RECOVER) Initiative, which seeks to understand, treat, and prevent the post-acute sequelae of SARS-CoV-2 infection (PASC). For more information on RECOVER, visit https://recovercovid.org/

We would like to thank the National Community Engagement Group (NCEG), all patient, caregiver and community Representatives, and all the participants enrolled in the RECOVER Initiative.

Finally, the authors gratefully acknowledge the contributions of Yong Chen, Xiaokang Liu, Miranda Higginbotham, and Alexander Shorrock.

## REFERENCES

[1] Huerne K, Filion KB, Grad R, Ernst P, Gershon AS, Eisenberg MJ. Epidemiological and clinical perspectives of long COVID syndrome. Am J Med Open 2023;9:100033. 10.1016/j.ajmo.2023.100033.

[2] Zheng Y-B, Zeng N, Yuan K, Tian S-S, Yang Y-B, Gao N, et al. Prevalence and risk factor for long COVID in children and adolescents: A meta-analysis and systematic review. J Infect Public Health 2023;16:660–72. 10.1016/j.jiph.2023.03.005.

[3] Rao S, Gross RS, Mohandas S, Stein CR, Case A, Dreyer B, et al. Postacute Sequelae of SARS-CoV-2 in Children. Pediatrics 2024;153:e2023062570. 10.1542/peds.2023-062570.

[4] Rao S, Lee GM, Razzaghi H, Lorman V, Mejias A, Pajor NM, et al. Clinical features and burden of post-acute sequelae of SARS-CoV-2 infection in children and adolescents: an exploratory EHR-based cohort study from the RECOVER program. MedRxiv Prepr Serv Health Sci 2022:2022.05.24.22275544. 10.1101/2022.05.24.22275544.

[5] Lorman V, Rao S, Jhaveri R, Case A, Mejias A, Pajor NM, et al. Understanding pediatric long COVID using a tree-based scan statistic approach: An EHR-based cohort study from the RECOVER Program. JAMIA Open 2023:ooad016. 10.1093/jamiaopen/ooad016.

[6] Borch L, Holm M, Knudsen M, Ellermann-Eriksen S, Hagstroem S. Long COVID symptoms and duration in SARS-CoV-2 positive children - a nationwide cohort study. Eur J Pediatr 2022;181:1597–607. 10.1007/s00431-021-04345-z.

[7] Fainardi V, Meoli A, Chiopris G, Motta M, Skenderaj K, Grandinetti R, et al. Long COVID in Children and Adolescents. Life Basel Switz 2022;12:285. 10.3390/life12020285.

[8] Radtke T, Ulyte A, Puhan MA, Kriemler S. Long-term Symptoms After SARS-CoV-2 Infection in Children and Adolescents. JAMA 2021;326:869–71. 10.1001/jama.2021.11880.

[9] Thallapureddy K, Thallapureddy K, Zerda E, Suresh N, Kamat D, Rajasekaran K, et al. Long-Term Complications of COVID-19 Infection in Adolescents and Children. Curr Pediatr Rep 2022;10:11–7. 10.1007/s40124-021-00260-x.

[10] Pellegrino R, Chiappini E, Licari A, Galli L, Marseglia GL. Prevalence and clinical presentation of long COVID in children: a systematic review. Eur J Pediatr 2022;181:3995–4009. 10.1007/s00431-022-04600-x.

[11] Reese JT, Blau H, Bergquist T, Loomba JJ, Callahan T, Laraway B, et al. Generalizable Long COVID Subtypes: Findings from the NIH N3C and RECOVER Programs. Infectious Diseases (except HIV/AIDS); 2022. 10.1101/2022.05.24.22275398.

[12] Zhang H, Zang C, Xu Z, Zhang Y, Xu J, Bian J, et al. Data-driven identification of post-acute SARS-CoV-2 infection subphenotypes. Nat Med 2022. 10.1038/s41591-022-02116-3.

[13] Information for Healthcare Providers about Multisystem Inflammatory Syndrome in Children (MIS-C) n.d.

[14] Rao S, Jing N, Liu X, Lorman V, Maltenfort M, Schuchard J, et al. Clinical Subphenotypes of Multisystem Inflammatory Syndrome in Children: An EHR-based cohort study from the RECOVER program. Pediatrics; 2022. 10.1101/2022.09.26.22280364.

[15] Botdorf M, Dickinson K, Lorman V, Razzaghi H, Marchesani N, Rao S, et al. EHR-based Case Identification of Pediatric Long COVID: A Report from the RECOVER EHR Cohort 2024. 10.1101/2024.05.23.24307492.

[16] De Freitas JK, Johnson KW, Golden E, Nadkarni GN, Dudley JT, Bottinger EP, et al. Phe2vec: Automated disease phenotyping based on unsupervised embeddings from electronic health records. Patterns N Y N 2021;2:100337. 10.1016/j.patter.2021.100337.

[17] Rao S, Lee GM, Razzaghi H, Lorman V, Mejias A, Pajor NM, et al. Clinical Features and Burden of Postacute Sequelae of SARS-CoV-2 Infection in Children and Adolescents. JAMA Pediatr 2022;176:1000. 10.1001/jamapediatrics.2022.2800.

[18] Lopez-Leon S, Wegman-Ostrosky T, Ayuzo Del Valle NC, Perelman C, Sepulveda R, Rebolledo PA, et al. Long-COVID in children and adolescents: a systematic review and meta-analyses. Sci Rep 2022;12:9950. 10.1038/s41598-022-13495-5.

[19] Jason LA, Jordan K, Miike T, Bell DS, Lapp C, Torres-Harding S, et al. A Pediatric Case Definition for Myalgic Encephalomyelitis and Chronic Fatigue Syndrome. J Chronic Fatigue Syndr 2006;13:1–44. 10.1300/J092v13n02_01.

[20] Solomon L, Reeves WC. Factors Influencing the Diagnosis of Chronic Fatigue Syndrome. Arch Intern Med 2004;164:2241. 10.1001/archinte.164.20.2241.

[21] Valdez AR, Hancock EE, Adebayo S, Kiernicki DJ, Proskauer D, Attewell JR, et al. Estimating Prevalence, Demographics, and Costs of ME/CFS Using Large Scale Medical Claims Data and Machine Learning. Front Pediatr 2019;6:412. 10.3389/fped.2018.00412.

[22] Bakken IJ, Tveito K, Gunnes N, Ghaderi S, Stoltenberg C, Trogstad L, et al. Two age peaks in the incidence of chronic fatigue syndrome/myalgic encephalomyelitis: a population-based registry study from Norway 2008-2012. BMC Med 2014;12:167. 10.1186/s12916-014-0167-5.

[23] Beyond Myalgic Encephalomyelitis/Chronic Fatigue Syndrome: Redefining an Illness. Washington, D.C.: National Academies Press; 2015. 10.17226/19012.

[24] Gross RS, Thaweethai T, Kleinman LC, Snowden JN, Rosenzweig EB, Milner JD, et al. Characterizing Long COVID in Children and Adolescents. JAMA 2024. 10.1001/jama.2024.12747.

[25] Khullar D, Zhang Y, Zang C, Xu Z, Wang F, Weiner MG, et al. Racial/Ethnic Disparities in Post-acute Sequelae of SARS-CoV-2 Infection in New York: an EHR-Based Cohort Study from the RECOVER Program. J Gen Intern Med 2023;38:1127–36. 10.1007/s11606-022-07997-1.

[26] Simon TD, Cawthon ML, Popalisky J, Mangione-Smith R. Development and Validation of the Pediatric Medical Complexity Algorithm (PMCA) Version 2.0. Hosp Pediatr 2017;7:373–7. 10.1542/hpeds.2016-0173.

[27] Mikolov T, Sutskever I, Chen K, Corrado G, Dean J. Distributed Representations of Words and Phrases and their Compositionality 2013. 10.48550/ARXIV.1310.4546.

[28] Arora S, Liang Y, Ma T. A simple but tough-to-beat baseline for sentence embeddings. 5th Int Conf Learn Represent ICLR 2017 2019.

[29] McInnes L, Healy J, Saul N, Großberger L. UMAP: Uniform Manifold Approximation and Projection. J Open Source Softw 2018;3:861. 10.21105/joss.00861.

[30] McInnes L, Healy J, Astels S. hdbscan: Hierarchical density based clustering. J Open Source Softw 2017;2:205. 10.21105/joss.00205.

[31] Forrest CB, Burrows EK, Mejias A, Razzaghi H, Christakis D, Jhaveri R, et al. Severity of Acute COVID-19 in Children <18 Years Old March 2020 to December 2021. Pediatrics 2022;149:e2021055765. 10.1542/peds.2021-055765.

[32] Řehůřek R, Sojka P. Software Framework for Topic Modelling with Large Corpora. Proc. LREC 2010 Workshop New Chall. NLP Framew., Valletta, Malta: ELRA; 2010, p. 45–50.

[33] Ho DE, Imai K, King G, Stuart EA. MatchIt : Nonparametric Preprocessing for Parametric Causal Inference. J Stat Softw 2011;42. 10.18637/jss.v042.i08.

