## Supplementary Materials for "Pediatric Long COVID Subphenotypes: An EHR-based study from the RECOVER program"

**SUPPLEMENTARY METHODS**

**PASC Subphenotype pipeline**

In this section, we describe in greater detail the components of the pipeline and the process for choosing hyperparameters. The first two steps (up through the embedded vectors representing patients’ clinical histories during the post-acute period) are part of the Phe2Vec algorithm; steps 3-5 are an extension of the method for the purposes of subphenotype detection and are original to this study. Steps 2-4 were first run against Cohort A multiple times for different hyperparameter values described below and output was compared in step 5. Once the pipeline was finalized, steps 2-4 were run a final time on each of Cohort A and B.

1. *Concept embedding model*

The first step of our pipeline was to learn 200-dimensional Euclidean embeddings for concepts (corresponding to codes) from the diagnosis (ICD10CM), medication (Clinical Drug Forms in RxNorm), and procedure (ICD10PCS, HCPCS, CPT4) vocabularies. We used the skip-gram version of the Word2Vec algorithm with negative sampling, a window size of 5, and ignored all codes that occurred fewer than 3 times in our training corpus. The model was trained for 15 epochs.

To construct the training corpus, we started with 9,835,122 patients in the pediatric RECOVER database. For each calendar month and year combination, we identified all distinct codes from the specified vocabularies which occurred during the given period. For each such period, we then constructed a sentence by concatenating the words in random order. Filtering out sentences containing only a single word resulted in a corpus of 99,413,139 sentences across 9,168,152 patients on a combined vocabulary of 99,371 words. The trained model (ignoring codes occurring fewer than 3 times) produced embeddings for 77,337 codes.

2. *Patient clinical history embeddings*

The second step of our pipeline was to use the learned embeddings of codes to represent the post-acute clinical histories of patients in the PASC cohort. For each patient in the cohort, we identified the distinct codes from the specified vocabularies that occurred during the 28-179 day post-acute period. We assembled these into sentences by concatenate codes in random order. To embed these sentences, we used the Simple but Tough to Beat Baseline for Sentence Embeddings^1^ with weighting parameter a=0.001. The result of this process was, for each patient in the PASC cohort, a representation of the patient’s post-acute clinical history by a 200-dimensional vector.

3. *Dimensional reduction*

To avoid difficulties in clustering resulting from sparsity of our dataset represented in 200-dimensional space (the “curse of dimensionality”), we elected to embed vectors in two dimensions, both to facilitate clustering and enable simple visualization. We used the Uniform Manifold Approximation and Projection algorithm (UMAP)^2^. To run the UMAP algorithm, we chose a min_dist parameter of 0 to facilitate clustering. The n_neighbors parameter, which controls balance between local and global structure in constructing lower-dimensional embeddings, was tuned from a grid of multiples of 5 between 5 and 500 using the validation method described below. Once this parameter was finalized, the output of this step was, for each patient in the PASC cohort, a 2-dimensional vector representing their post-acute clinical history.

4. *Clustering*

Due to the irregular shape and non-uniform density of the embedded clinical history dataset, we used hierarchical density-based clustering, implemented by the HDBSCAN algorithm^3^. Clusters detected were required to contain at least 2% of the cohort. To minimize the number of unclustered patients, we opted for the least conservative implementation of HDBSCAN with a min_samples value of 1. The number of clusters is not pre-specified in HDBSCAN but rather emerges from the algorithm. The output of this stage was for each patient either a cluster assignment or an indicator that the patient was not clustered. In our main analysis, we assigned each patient unclustered by HDBSCAN to the cluster whose centroid was nearest to them; we included the original model output allows for unclustered patients as a sensitivity analysis.

5. *Hyperparameter selection*

In order to tune the n_neighbors hyperparameter described above, we ran the pipeline for each multiple of 5 between 5 and 500. Thus, each parameter value produced a unique clustering of the PASC cohort, resulting in 100 clusterings. To select the value of n_neighbors that produced the most clinically plausible clustering, we first restricted to values which resulted in no fewer than 5 and no more than 20 clusters with no more than 25% of the cohort remaining unclustered. Of the resulting clusterings, we simplified the landscape further by using variation of information (VI)^4^ to identify n_neighbors values that produced similar clusterings. VI is a metric on the set of clusterings of a given (fixed) dataset that measures the extent to which two clusterings assign patients to the same groups. Specifically, we calculated pairwise VI-distances between all remaining clusterings, then grouped them into four groups of like clusterings (using simple hierarchical clustering). From each group, we then selected the clustering (and corresponding n_neighbors value) which had the smallest proportion of unclustered patients. This reduced the set of n_neighbors values in consideration down to values of 15, 85, 95, and 300.

We then conducted descriptive analyses, producing analogs of Figure 3 and Tables 2-3 for each of the four remaining clusterings. While we found substantial overlap in the clinical profiles represented by the clusters across different values of n_neighbors, we selected the clustering corresponding to n_neighbors=85 as it represented respiratory/cardiac, gastrointestinal, musculoskeletal/pain, headache, fatigue, and neuropsychiatric subtypes with the smallest number of clusters, and there were no additional presentations identified in clusterings corresponding to other choices of the n_neigbors parameter.

**Matched control cohort**

To evaluate the extent to which clusters and subphenotypes identified in the Long COVID cohort were specific to Long COVID versus reflective of clinical profiles exhibited in more general care-seeking populations, we identified a control cohort of patients with no evidence of SARS-CoV-2 infection. We selected patients with visits recorded in the pediatric RECOVER EHR data during the same time period as in our Long COVID cohort. Index date was set to the date of a random visit during this time period. To ensure a comparable volume of post-index utilization as in the Long COVID cohort, we additionally required patients to have two visits at least 28 days apart during the 28-179 day post-index period. As in the Long COVID cohort, we excluded patients with evidence of existing complex chronic conditions in the 3 years prior to index date. Finally, we used 1:1 nearest neighbor propensity score matching to ensure our control cohort had a similar distribution of potential confounders to the Long COVID cohort. Variables used in the propensity score model included age group, sex, racial and ethnic group, time period of index date, and presence of existing (non-complex) chronic conditions across each of 17 body systems. After matching, all covariates were adequately balanced (absolute standardized mean differences less than 0.1).

**Figure 1S: Heatmap of incident post-acute diagnoses by subphenotype, Cohort A**

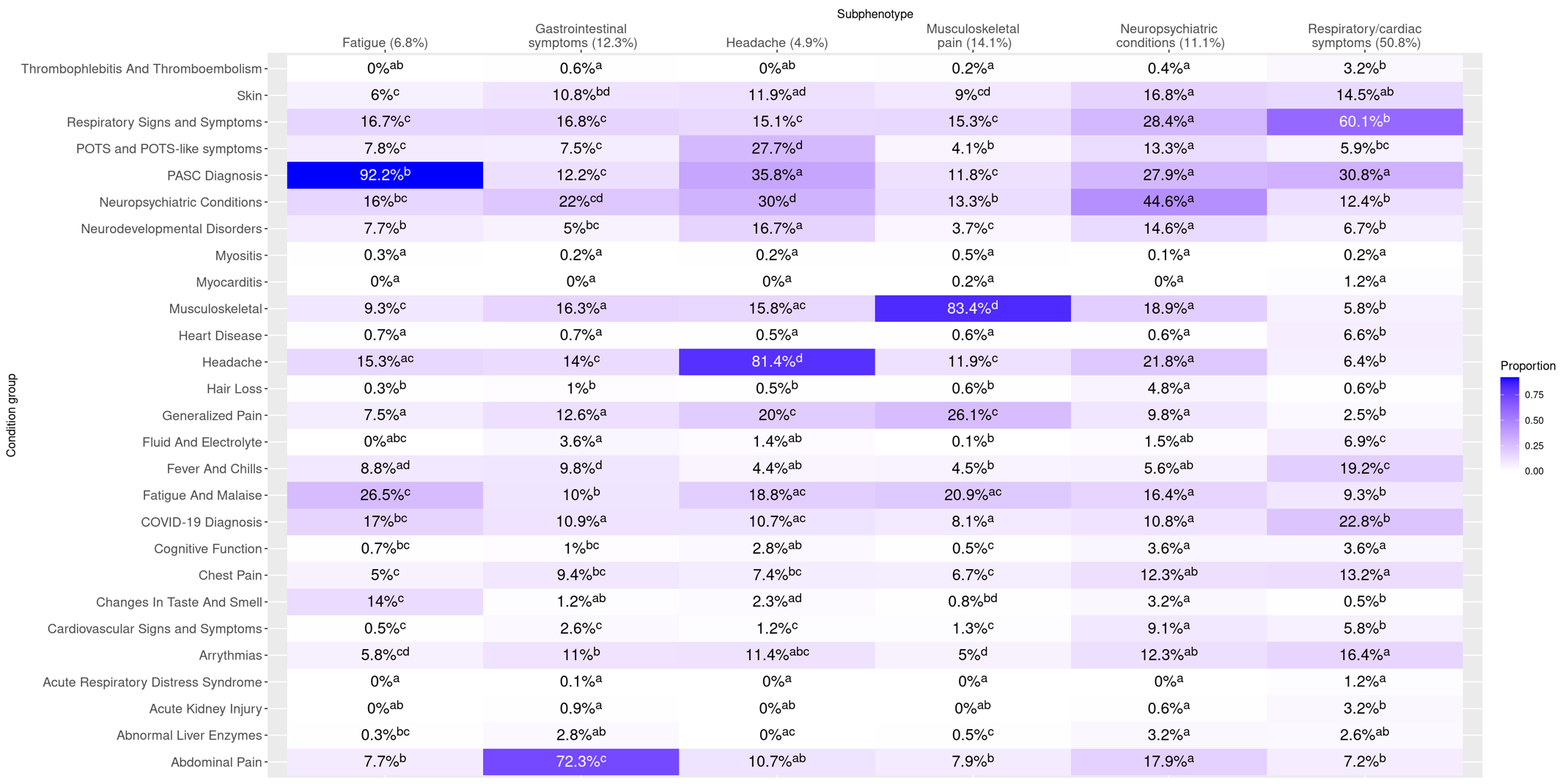

**Figure 2S-A: Heatmap of incident post-acute diagnoses by cluster, Cohort A**

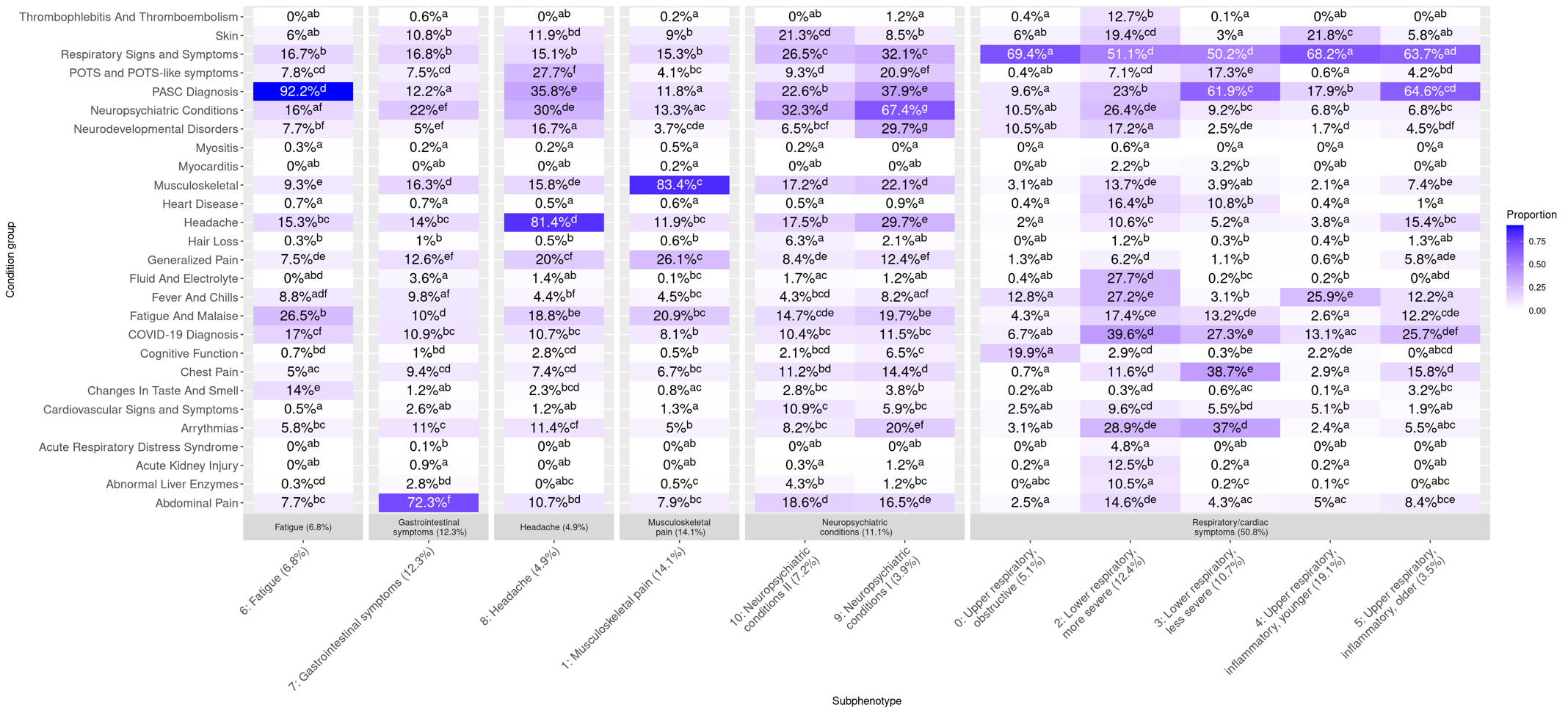
**Figure 2S-B: Heatmap of incident post-acute diagnoses by cluster, Cohort B**

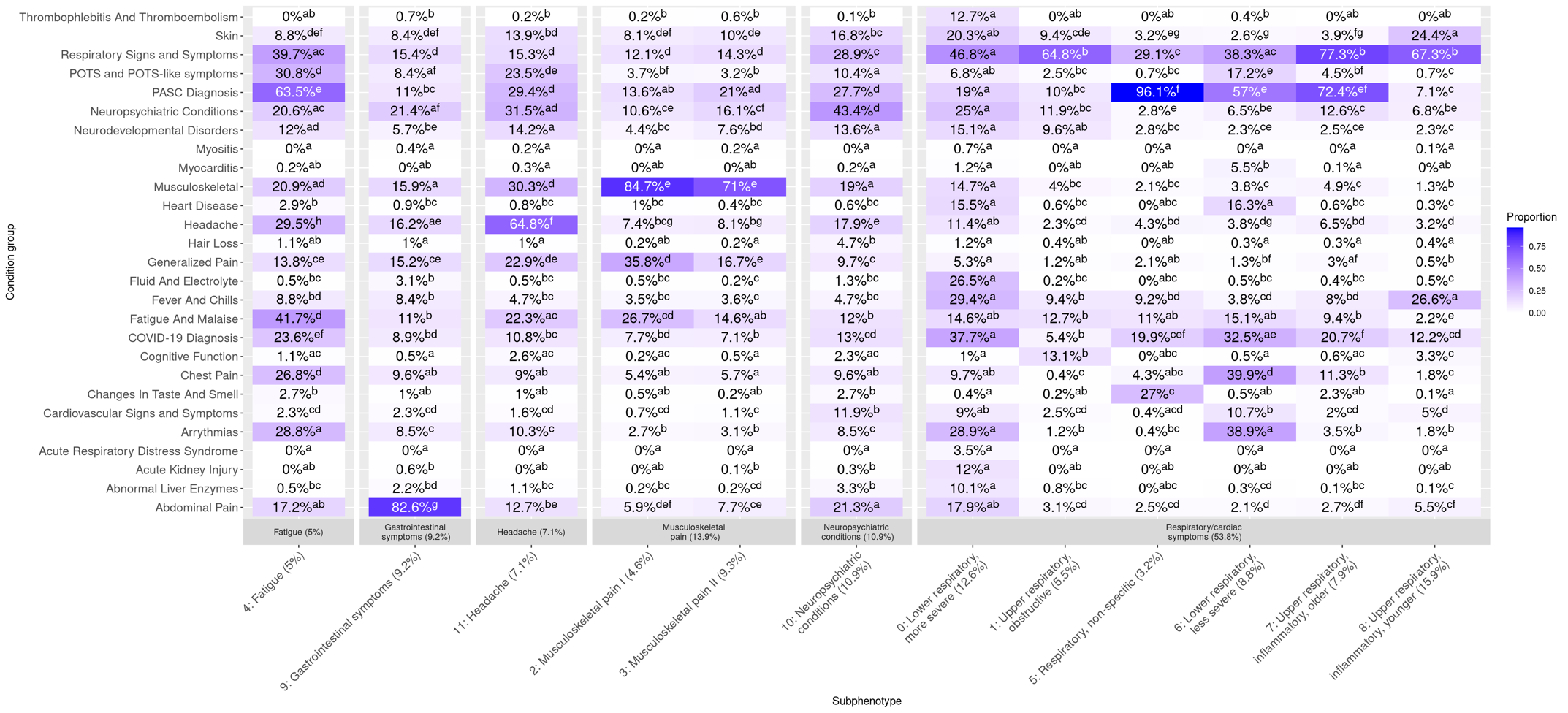

**Table 1S-A: Demographic and clinical characteristics of clusters, cohort A**

Note: cells marked with an asterisk have been modified by a random count between 0 and 4 to prevent reidentification of that cell or a cell in the same group.

| **Subphenotype** |  | Fatigue (600, 6.8%) | Gastrointestinal symptoms (1078, 12.3%) | Headache (430, 4.9%) | Musculoskeletal pain (1232, 14.1%) | Neuropsychiatric conditions (974, 11.1%) | | Respiratory/cardiac symptoms (4454, 50.8%) | | | | |
| --- | --- | --- | --- | --- | --- | --- | --- | --- | --- | --- | --- | --- |
| **Cluster** |  | 6: Fatigue (600, 6.8%) | 7: Gastrointestinal symptoms (1078, 12.3%) | 8: Headache (430, 4.9%) | 1: Musculoskeletal pain (1232, 14.1%) | 9: Neuropsychiatric conditions I (340, 3.9%) | 10: Neuropsychiatric conditions II (634, 7.2%) | 0: Upper respiratory, obstructive (447, 5.1%) | 2: Lower respiratory, more severe (1089, 12.4%) | 3: Lower respiratory, less severe (935, 10.7%) | 4: Upper respiratory, inflammatory, younger (1672, 19.1%) | 5: Upper respiratory, inflammatory, older (311, 3.5%) |
| **Age group (n/%)** | <1 | 5 (0.8%) | 13 (1.2%) | 0 (0%) | 16 (1.3%) | 0 (0%) | 0 (0%) | 72 (16.1%) | 258 (23.7%) | 18 (1.9%) | 488 (29.2%) | 2 (0.1%)* |
|  | 1-4 | 45 (7.5%) | 73 (6.8%) | 3 (0.7%)* | 58 (4.7%) | 5 (1.5%)* | 30 (4.7%) | 241 (53.9%) | 216 (19.8%) | 35 (3.7%) | 626 (37.4%) | 15 (4.8%)* |
|  | 5-11 | 157 (26.2%) | 280 (26%) | 96 (22.3%)* | 284 (23.1%) | 66 (19.4%)* | 142 (22.4%) | 104 (23.3%) | 154 (14.1%) | 164 (17.5%) | 378 (22.6%) | 75 (24.1%) |
|  | 12-15 | 185 (30.8%) | 275 (25.5%) | 154 (35.8%) | 410 (33.3%) | 104 (30.6%) | 194 (30.6%) | 13 (2.9%) | 160 (14.7%) | 302 (32.3%) | 93 (5.6%) | 73 (23.5%) |
|  | 16-20 | 208 (34.7%) | 437 (40.5%) | 177 (41.2%) | 464 (37.7%) | 165 (48.5%) | 268 (42.3%) | 17 (3.8%) | 301 (27.6%) | 416 (44.5%) | 87 (5.2%) | 146 (46.9%) |
| **Sex (n/%)** | Female | 306 (51%) | 722 (67%) | 296 (68.8%) | 661 (53.7%) | 203 (59.7%) | 422 (66.6%) | 179 (40%) | 546 (50.1%) | 511 (54.7%) | 762 (45.6%) | 169 (54.3%) |
|  | Male/Other/Unknown | 294 (49%) | 356 (33%) | 134 (31.2%) | 571 (46.3%) | 137 (40.3%) | 212 (33.4%) | 268 (60%) | 543 (49.9%) | 424 (45.3%) | 910 (54.4%) | 142 (45.7%) |
| **Race/ethnicity (n/%)** | Non-Hispanic Black/AA | 41 (6.8%) | 115 (10.7%) | 41 (9.5%) | 176 (14.3%) | 41 (12.1%) | 92 (14.5%) | 71 (15.9%) | 209 (19.2%) | 97 (10.4%) | 255 (15.3%) | 26 (8.4%) |
|  | Non-Hispanic Asian/PI | 14 (2.3%) | 27 (2.5%) | 12 (2.8%) | 36 (2.9%) | 7 (2.1%) | 28 (4.4%) | 12 (2.7%) | 49 (4.5%) | 22 (2.4%) | 93 (5.6%) | 14 (4.5%) |
|  | Hispanic | 97 (16.2%) | 291 (27%) | 67 (15.6%) | 216 (17.5%) | 52 (15.3%) | 211 (33.3%) | 97 (21.7%) | 292 (26.8%) | 157 (16.8%) | 452 (27%) | 73 (23.5%) |
|  | Non-Hispanic White | 372 (62%) | 579 (53.7%) | 270 (62.8%) | 688 (55.8%) | 204 (60%) | 246 (38.8%) | 220 (49.2%) | 439 (40.3%) | 543 (58.1%) | 637 (38.1%) | 174 (55.9%) |
|  | Multiple | 3 (0.5%)* | 12 (1.1%) | 8 (1.9%) | 38 (3.1%) | 9 (2.6%) | 1 (0.2%)* | 13 (2.9%) | 25 (2.3%) | 27 (2.9%) | 47 (2.8%) | 0 (0%) |
|  | Other/Unknown | 73 (12.2%)* | 54 (5%) | 32 (7.4%) | 78 (6.3%) | 27 (7.9%) | 56 (8.8%)* | 34 (7.6%) | 75 (6.9%) | 89 (9.5%) | 188 (11.2%) | 24 (7.7%) |
| **Cohort entry period (n/%)** | Mar-Jun 2020 | 15 (2.5%) | 26 (2.4%) | 6 (1.4%) | 19 (1.5%) | 8 (2.4%) | 15 (2.4%) | 6 (1.3%) | 46 (4.2%) | 13 (1.4%) | 10 (0.6%) | 4 (1.3%)* |
|  | Jul-Oct 2020 | 22 (3.7%) | 70 (6.5%) | 24 (5.6%) | 106 (8.6%) | 24 (7.1%) | 42 (6.6%) | 13 (2.9%) | 81 (7.4%) | 56 (6%) | 45 (2.7%) | 8 (2.6%)* |
|  | Nov-Feb 2021 | 53 (8.8%) | 175 (16.2%) | 53 (12.3%) | 228 (18.5%) | 46 (13.5%) | 103 (16.2%) | 49 (11%) | 119 (10.9%) | 141 (15.1%) | 108 (6.5%) | 17 (5.5%) |
|  | Mar-Jun 2021 | 86 (14.3%) | 60 (5.6%) | 26 (6%) | 83 (6.7%) | 16 (4.7%) | 37 (5.8%) | 20 (4.5%) | 125 (11.5%) | 89 (9.5%) | 73 (4.4%) | 7 (2.3%) |
|  | Jul-Oct 2021 | 104 (17.3%) | 182 (16.9%) | 71 (16.5%) | 215 (17.5%) | 72 (21.2%) | 96 (15.1%) | 55 (12.3%) | 184 (16.9%) | 182 (19.5%) | 236 (14.1%) | 80 (25.7%) |
|  | Nov-Feb 2022 | 188 (31.3%) | 389 (36.1%) | 151 (35.1%) | 427 (34.7%) | 116 (34.1%) | 234 (36.9%) | 205 (45.9%) | 303 (27.8%) | 284 (30.4%) | 692 (41.4%) | 122 (39.2%) |
|  | Mar-Jun 2022 | 95 (15.8%) | 107 (9.9%) | 58 (13.5%) | 86 (7%) | 37 (10.9%) | 72 (11.4%) | 58 (13%) | 134 (12.3%) | 110 (11.8%) | 285 (17%) | 34 (10.9%) |
|  | Jul-Aug 2022 | 37 (6.2%) | 69 (6.4%) | 41 (9.5%) | 68 (5.5%) | 21 (6.2%) | 35 (5.5%) | 41 (9.2%) | 97 (8.9%) | 60 (6.4%) | 223 (13.3%) | 39 (12.5%) |
| **ICU (acute) (n/%)** |  | 0 (0%) | 10 (0.9%) | 0 (0%) | 7 (0.6%) | 2 (0.6%)* | 9 (1.4%) | 3 (0.7%)* | 95 (8.7%) | 14 (1.5%) | 7 (0.4%) | 0 (0%) |
| **Hospitalization (acute) (n/%)** |  | 1 (0.2%)* | 82 (7.6%) | 10 (2.3%) | 52 (4.2%) | 22 (6.5%) | 14 (2.2%) | 26 (5.8%) | 302 (27.7%) | 30 (3.2%) | 57 (3.4%) | 3 (1.0%)* |
| **COVID acute phase severity of illness (n/%)** | Asymptomatic | 525 (87.5%) | 572 (53.1%) | 270 (62.8%) | 752 (61%) | 235 (69.1%) | 403 (63.6%) | 259 (57.9%) | 662 (60.8%) | 733 (78.4%) | 881 (52.7%) | 232 (74.6%) |
|  | Mild | 66 (11%) | 405 (37.6%) | 136 (31.6%) | 417 (33.8%) | 91 (26.8%) | 208 (32.8%) | 143 (32%) | 184 (16.9%) | 149 (15.9%) | 671 (40.1%) | 70 (22.5%) |
|  | Moderate | 8 (1.3%)* | 82 (7.6%) | 16 (3.7%) | 51 (4.1%) | 9 (2.6%) | 15 (2.4%) | 30 (6.7%) | 101 (9.3%) | 19 (2%) | 105 (6.3%) | 9 (2.9%) |
|  | Severe | 1 (0.2%)* | 19 (1.8%) | 8 (1.9%) | 12 (1%) | 5 (1.5%) | 8 (1.3%) | 15 (3.4%) | 142 (13%) | 34 (3.6%) | 15 (0.9%) | 0 (0%) |
| **Presence of existing chronic condition (n/%)** |  | 172 (28.7%) | 496 (46%) | 156 (36.3%) | 445 (36.1%) | 178 (52.4%) | 283 (44.6%) | 189 (42.3%) | 368 (33.8%) | 270 (28.9%) | 556 (33.3%) | 113 (36.3%) |
| **Most common diagnoses** |  | U09.9: Post COVID-19 condition, unspecified (69.7%)  B94.8: Sequelae of other specified infectious and parasitic diseases (27.7%) | R10.9: Unspecified abdominal pain (39.0%)  R10.84: Generalized abdominal pain (26.4%) | R51.9: Headache, unspecified (47.0%)  U09.9: Post COVID-19 condition, unspecified (32.3%) | G89.29: Other chronic pain (22.6%) | U09.9: Post COVID-19 condition, unspecified (30.9%)  F41.9: Anxiety disorder, unspecified (23.2%) | U09.9: Post COVID-19 condition, unspecified (20.5%) | R06.83: Snoring (40.9%)  R09.81: Nasal congestion (25.3%)  G47.30: Sleep apnea, unspecified (21.5%) | U07.1: Emergency use of U07.1 \| COVID-19 (38.8%)  R50.9: Fever, unspecified (22.2%) | U09.9: Post COVID-19 condition, unspecified (52.9%)  R07.9: Chest pain, unspecified (29.6%)  R06.02: Shortness of breath (29.4%) U07.1: Emergency use of U07.1 \| COVID-19 (27.3%)  R00.2: Palpitations (20.1%) | R05.9: Cough, unspecified (34.03%)  R50.9: Fever, unspecified (24.2%)  R09.81: Nasal congestion (21.2%) | U09.9: Post COVID-19 condition, unspecified (62.1%)  R05.9: Cough, unspecified (30.9%)  U07.1: Emergency use of U07.1 \| COVID-19 (25.4%) |

**Table 1S-B: Demographic and clinical characteristics of clusters, cohort B**

Note: cells marked with an asterisk have been modified by a random count between 0 and 4 to prevent reidentification of that cell or a cell in the same group.

| **Subphenotype** |  | Fatigue (441, 5%) | Gastrointestinal symptoms (810, 9.2%) | Headache (620, 7.1%) | Musculoskeletal pain (1218, 13.9%) | | Neuropsychiatric conditions (952, 10.9%) | Respiratory/cardiac symptoms (4716, 53.8%) | | | | | |
| --- | --- | --- | --- | --- | --- | --- | --- | --- | --- | --- | --- | --- | --- |
| **Cluster** |  | 4: Fatigue (441, 5%) | 9: Gastrointestinal symptoms (810, 9.2%) | 11: Headache (620, 7.1%) | 2: Musculoskeletal pain I (405, 4.6%) | 3: Musculoskeletal pain II (813, 9.3%) | 10: Neuropsychiatric conditions (952, 10.9%) | 0: Lower respiratory, more severe (1106, 12.6%) | 1: Upper respiratory, obstructive (480, 5.5%) | 5: Respiratory, non-specific (282, 3.2%) | 6: Lower respiratory, less severe (767, 8.8%) | 7: Upper respiratory, inflammatory, older (691, 7.9%) | 8: Upper respiratory, inflammatory, younger (1390, 15.9%) |
| **Age group (n/%)** | <1 | 0 (0%) | 6 (0.7%) | 0 (0%) | 10 (2.5%) | 7 (0.9%) | 4 (0.4%)* | 254 (23%) | 94 (19.6%) | 9 (3.2%) | 20 (2.6%) | 11 (1.6%) | 428 (30.8%) |
|  | 1-4 | 3 (0.7%)* | 49 (6%) | 16 (2.6%) | 11 (2.7%) | 49 (6%) | 21 (2.2%)* | 197 (17.8%) | 239 (49.8%) | 29 (10.3%) | 31 (4%) | 71 (10.3%) | 595 (42.8%) |
|  | 5-11 | 89 (20.2%)* | 233 (28.8%) | 116 (18.7%) | 78 (19.3%) | 237 (29.2%) | 190 (20%) | 182 (16.5%) | 95 (19.8%) | 69 (24.5%) | 144 (18.8%) | 185 (26.8%) | 303 (21.8%) |
|  | 12-15 | 145 (32.9%) | 206 (25.4%) | 233 (37.6%) | 156 (38.5%) | 263 (32.3%) | 298 (31.3%) | 160 (14.5%) | 35 (7.3%) | 88 (31.2%) | 249 (32.5%) | 175 (25.3%) | 39 (2.8%) |
|  | 16-20 | 204 (46.3%) | 316 (39%) | 255 (41.1%) | 150 (37%) | 257 (31.6%) | 439 (46.1%) | 313 (28.3%) | 17 (3.5%) | 87 (30.9%) | 323 (42.1%) | 249 (36%) | 25 (1.8%) |
| **Sex (n/%)** | Female | 285 (64.6%) | 536 (66.2%) | 421 (67.9%) | 225 (55.6%) | 385 (47.4%) | 643 (67.5%) | 576 (52.1%) | 201 (41.9%) | 149 (52.8%) | 383 (49.9%) | 367 (53.1%) | 607 (43.7%) |
|  | Male/Other/Unknown | 156 (35.4%) | 274 (33.8%) | 199 (32.1%) | 180 (44.4%) | 428 (52.6%) | 309 (32.5%) | 530 (47.9%) | 279 (58.1%) | 133 (47.2%) | 384 (50.1%) | 324 (46.9%) | 783 (56.3%) |
| **Race/ethnicity (n/%)** | Non-Hispanic Black/AA | 43 (9.8%) | 105 (13%) | 73 (11.8%) | 52 (12.8%) | 125 (15.4%) | 139 (14.6%) | 197 (17.8%) | 90 (18.8%) | 30 (10.6%) | 76 (9.9%) | 67 (9.7%) | 233 (16.8%) |
|  | Non-Hispanic Asian/PI | 11 (2.5%) | 19 (2.3%) | 6 (1%) | 9 (2.2%) | 13 (1.6%) | 29 (3%) | 45 (4.1%) | 16 (3.3%) | 7 (2.5%) | 18 (2.3%) | 19 (2.7%) | 64 (4.6%) |
|  | Hispanic | 69 (15.6%) | 183 (22.6%) | 100 (16.1%) | 66 (16.3%) | 127 (15.6%) | 251 (26.4%) | 305 (27.6%) | 102 (21.2%) | 59 (20.9%) | 129 (16.8%) | 150 (21.7%) | 410 (29.5%) |
|  | Non-Hispanic White | 264 (59.9%) | 426 (52.6%) | 368 (59.4%) | 243 (60%) | 467 (57.4%) | 457 (48%) | 447 (40.4%) | 236 (49.2%) | 145 (51.4%) | 446 (58.1%) | 363 (52.5%) | 502 (36.1%) |
|  | Multiple | 9 (2%) | 18 (2.2%) | 16 (2.6%) | 11 (2.7%) | 23 (2.8%) | 15 (1.6%) | 27 (2.4%) | 16 (3.3%) | 6 (2.1%) | 25 (3.3%) | 12 (1.7%) | 27 (1.9%) |
|  | Other/Unknown | 45 (10.2%) | 59 (7.3%) | 57 (9.2%) | 24 (5.9%) | 58 (7.1%) | 61 (6.4%) | 85 (7.7%) | 20 (4.2%) | 35 (12.4%) | 73 (9.5%) | 80 (11.6%) | 154 (11.1%) |
| **Cohort entry period (n/%)** | Mar-Jun 2020 | 2 (0.7%)* | 21 (2.6%) | 10 (1.6%) | 7 (1.7%) | 37 (4.6%) | 19 (2%) | 50 (4.5%) | 12 (2.5%) | 2 (0.7%)* | 19 (2.5%) | 3 (0.4%) | 7 (0.5%) |
|  | Jul-Oct 2020 | 19 (4.3%)* | 50 (6.2%) | 38 (6.1%) | 22 (5.4%) | 86 (10.6%) | 52 (5.5%) | 71 (6.4%) | 18 (3.8%) | 1 (0.4%)* | 38 (5%) | 14 (2%)* | 27 (1.9%) |
|  | Nov-Feb 2021 | 47 (10.7%) | 148 (18.3%) | 102 (16.5%) | 79 (19.5%) | 160 (19.7%) | 153 (16.1%) | 170 (15.4%) | 55 (11.5%) | 12 (4.3%) | 120 (15.6%) | 40 (5.8%) | 111 (8%) |
|  | Mar-Jun 2021 | 33 (7.5%) | 50 (6.2%) | 43 (6.9%) | 27 (6.7%) | 87 (10.7%) | 62 (6.5%) | 120 (10.8%) | 27 (5.6%) | 13 (4.6%) | 91 (11.9%) | 29 (4.2%) | 57 (4.1%) |
|  | Jul-Oct 2021 | 79 (17.9%) | 113 (14%) | 102 (16.5%) | 67 (16.5%) | 105 (12.9%) | 147 (15.4%) | 162 (14.6%) | 53 (11%) | 77 (27.3%) | 138 (18%) | 132 (19.1%) | 199 (14.3%) |
|  | Nov-Feb 2022 | 155 (35.1%) | 295 (36.4%) | 220 (35.5%) | 143 (35.3%) | 247 (30.4%) | 346 (36.3%) | 304 (27.5%) | 213 (44.4%) | 104 (36.9%) | 235 (30.6%) | 255 (36.9%) | 567 (40.8%) |
|  | Mar-Jun 2022 | 59 (13.4%) | 79 (9.8%) | 59 (9.5%) | 30 (7.4%) | 59 (7.3%) | 112 (11.8%) | 124 (11.2%) | 58 (12.1%) | 55 (19.5%) | 86 (11.2%) | 138 (20%) | 230 (16.5%) |
|  | Jul-Aug 2022 | 47 (10.7%) | 54 (6.7%) | 46 (7.4%) | 30 (7.4%) | 32 (3.9%) | 61 (6.4%) | 105 (9.5%) | 44 (9.2%) | 18 (6.4%) | 40 (5.2%) | 80 (11.6%) | 192 (13.8%) |
| **ICU (acute) (n/%)** |  | 0 (0%) | 2 (0.2%)* | 5 (0.8%) | 4 (1.0%)* | 6 (0.7%) | 6 (0.6%) | 118 (10.7%) | 3 (0.6%)* | 0 (0%) | 13 (1.7%) | 3 (0.4%)* | 1 (0.1%)* |
| **Hospitalization (acute) (n/%)** |  | 4 (0.9%)* | 56 (6.9%) | 21 (3.4%) | 7 (1.7%) | 33 (4.1%) | 34 (3.6%) | 303 (27.4%) | 33 (6.9%) | 0 (0%) | 40 (5.2%) | 7 (1%) | 42 (3%) |
| **COVID acute phase severity of illness (n/%)** | Asymptomatic | 336 (76.2%) | 451 (55.7%) | 404 (65.2%) | 261 (64.4%) | 499 (61.4%) | 624 (65.5%) | 654 (59.1%) | 261 (54.4%) | 256 (90.8%) | 584 (76.1%) | 564 (81.6%) | 675 (48.6%) |
|  | Mild | 89 (20.2%) | 287 (35.4%) | 184 (29.7%) | 123 (30.4%) | 269 (33.1%) | 290 (30.5%) | 177 (16%) | 173 (36%) | 23 (8.2%)* | 130 (16.9%) | 113 (16.4%) | 607 (43.7%) |
|  | Moderate | 11 (2.5%) | 62 (7.7%) | 23 (3.7%) | 16 (4%) | 33 (4.1%) | 28 (2.9%) | 89 (8%) | 39 (8.1%) | 3 (1.1%)* | 18 (2.3%) | 9 (1.3%) | 89 (6.4%) |
|  | Severe | 5 (1.1%) | 10 (1.2%) | 9 (1.5%) | 5 (1.2%) | 12 (1.5%) | 10 (1.1%) | 186 (16.8%) | 7 (1.5%) | 0 (0%) | 35 (4.6%) | 5 (0.7%) | 19 (1.4%) |
| **Presence of existing chronic condition (n/%)** |  | 160 (36.3%) | 373 (46%) | 250 (40.3%) | 161 (39.8%) | 297 (36.5%) | 428 (45%) | 384 (34.7%) | 221 (46%) | 81 (28.7%) | 198 (25.8%) | 250 (36.2%) | 462 (33.2%) |
| **Most common diagnoses** |  | U09.9: Post COVID-19 condition, unspecified (57.6%)  R53.83: Other fatigue (30.8%)  U07.1: Emergency use of U07.1 \| COVID-19 (23.4%) R51.9: Headache, unspecified (23.1%)  R42: Dizziness and giddiness (22.2%) | R10.9: Unspecified abdominal pain (45.6%)  R10.84: Generalized abdominal pain (28.8%)  R10.13: Epigastric pain (21.2%)  K59.00: Constipation, unspecified (21.2%) | R51.9: Headache, unspecified (37.9%)  U09.9: Post COVID-19 condition, unspecified (25.5%) | G89.29: Other chronic pain (32.4%)  M62.81: Muscle weakness (generalized) (20.5%)  M54.50: Low back pain, unspecified (20.0%) |  | U09.9: Post COVID-19 condition, unspecified (23.9%) | U07.1: Emergency use of U07.1 \| COVID-19 (36.9%)  R50.9: Fever, unspecified (25.4%) | R06.83: Snoring (36.3%)  R09.81: Nasal congestion (21.5%) | U09.9: Post COVID-19 condition, unspecified (92.6%) | U09.9: Post COVID-19 condition, unspecified (46.2%)  U07.1: Emergency use of U07.1 \| COVID-19 (32.5%)  R07.9: Chest pain, unspecified (31.0%)  R06.02: Shortness of breath (23.2%)  B94.8: Sequelae of other specified infectious and parasitic diseases (20.6%) | U09.9: Post COVID-19 condition, unspecified (70.0%)  R05.3: Chronic cough (30.97%)  R05.9: Cough, unspecified (26.0%)  U07.1: Emergency use of U07.1 \| COVID-19 (20.7%) | R05.9: Cough, unspecified (33.2%)  R50.9: Fever, unspecified (25.2%)  R09.81: Nasal congestion (21.7%) |

**Figure 3S: Heatmap of incident follow-up diagnoses for matched control cohort**

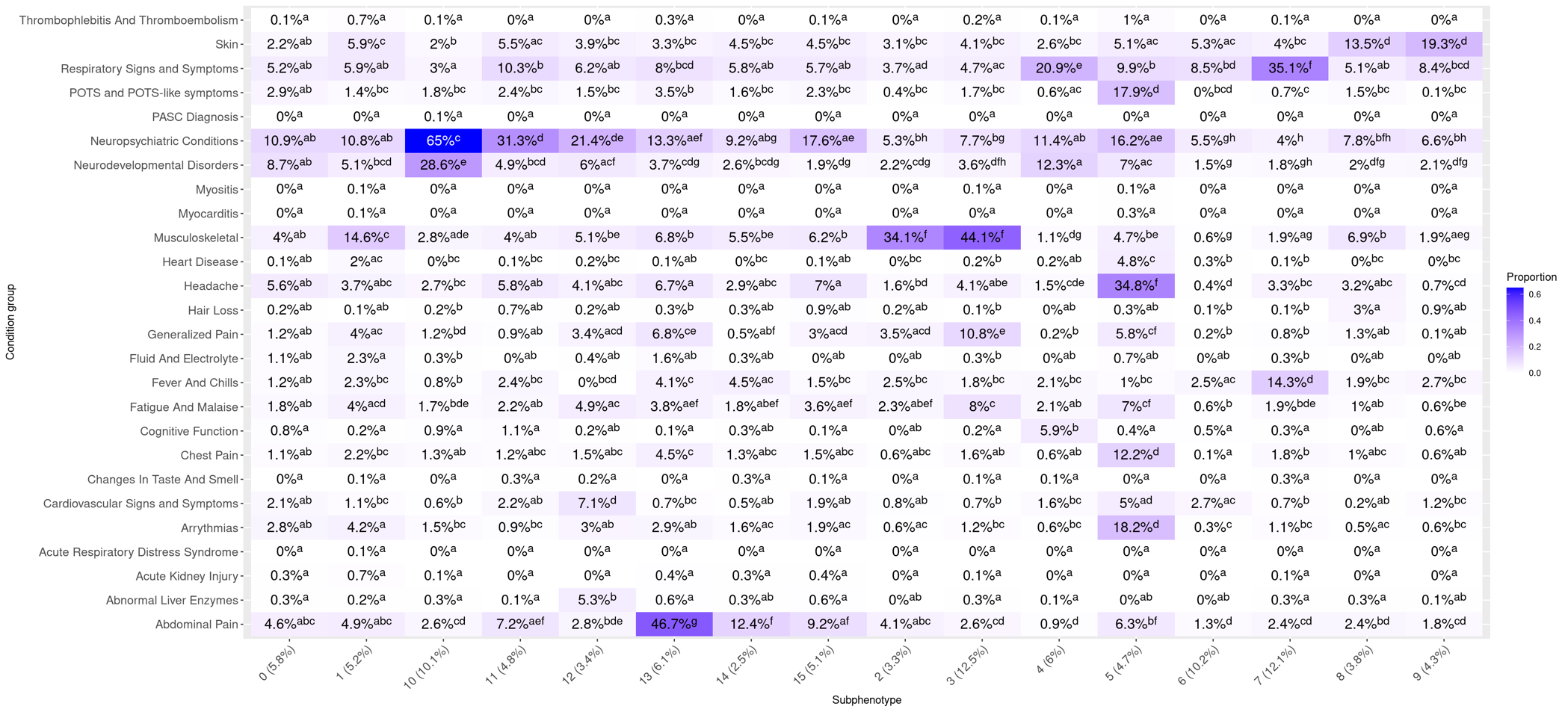

**Figure 4S: Subphenotype centroids: comparison of cohort A, cohort B, and matched control cohort.**

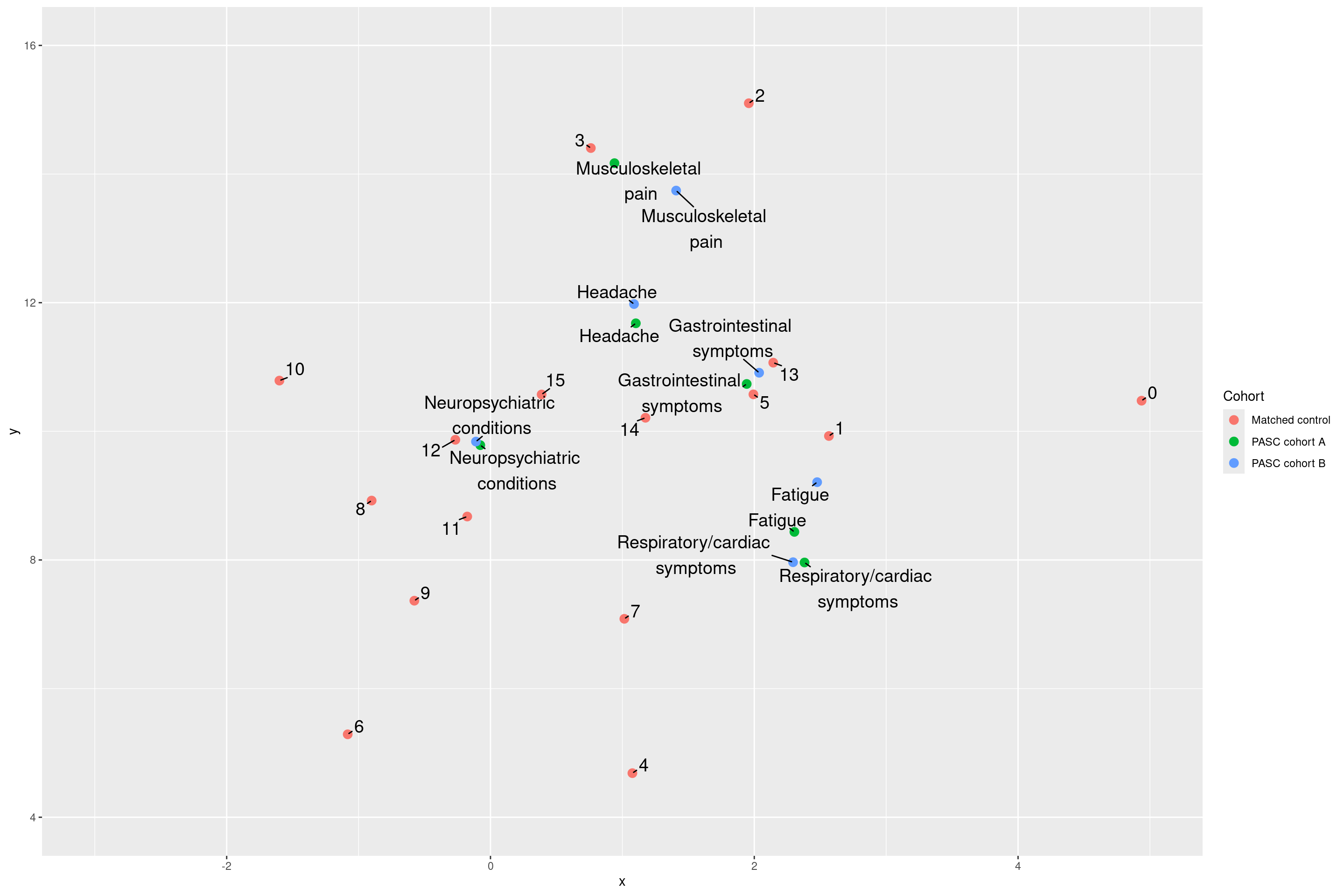
**Table 2S: Most common medications and procedures by subphenotype, cohort B**

| Subphenotype | Fatigue | Gastrointestinal symptoms | Headache | Musculoskeletal pain | Neuropsychiatric conditions | Respiratory/cardiac symptoms |
| --- | --- | --- | --- | --- | --- | --- |
| Most common procedures | 93005: Electrocardiogram, routine ECG with at least 12 leads; tracing only, w… (24.3%)  93010: Electrocardiogram, routine ECG with at least 12 leads; interpretation … (21.1%)  36415: Collection of venous blood by venipuncture (16.3%)  71046: Radiologic examination, chest; 2 views (14.7%)  93306: Echocardiography, transthoracic, real-time with image documentation (2… (14.3%) | 43239: Esophagogastroduodenoscopy, flexible, transoral; with biopsy, single o… (20.9%)  36415: Collection of venous blood by venipuncture (18.0%)  74018: Radiologic examination, abdomen; 1 view (16.1%)  76705: Ultrasound, abdominal, real time with image documentation; limited (eg… (15.1%)  76700: Ultrasound, abdominal, real time with image documentation; complete (11.4%) | 36415: Collection of venous blood by venipuncture (15.3%)  70551: Magnetic resonance (eg, proton) imaging, brain (including brain stem);… (14.7%)  97110: Therapeutic procedure, 1 or more areas, each 15 minutes; therapeutic e… (11.1%)  93005: Electrocardiogram, routine ECG with at least 12 leads; tracing only, w… (10.8%) | 97110: Therapeutic procedure, 1 or more areas, each 15 minutes; therapeutic e… (27.3%)  97161: Physical therapy evaluation: low complexity, requiring these component… (20.1%)  97112: Therapeutic procedure, 1 or more areas, each 15 minutes; neuromuscular… (12.2%) 36415: Collection of venous blood by venipuncture (10.2%) | 36415: Collection of venous blood by venipuncture (14.2%) | 71046: Radiologic examination, chest; 2 views (17.5%)  93005: Electrocardiogram, routine ECG with at least 12 leads; tracing only, w… (17.3%)  93010: Electrocardiogram, routine ECG with at least 12 leads; interpretation … (16.4%)  36415: Collection of venous blood by venipuncture (11.4%)  93306: Echocardiography, transthoracic, real-time with image documentation (2… (11.4%) |
| Most common medications | 745678: albuterol Metered Dose Inhaler (11.6%) | 373149: ondansetron Disintegrating Oral Tablet (21.4%)  876077: calcium chloride / lactate / potassium chloride / sodium chloride Inje… (20.4%)  1870358: polyethylene glycol 3350 Powder for Oral Solution (19.3%)  376327: ondansetron Injectable Solution (18.9%)  378236: omeprazole Delayed Release Oral Capsule (17.2%) | 370674: ibuprofen Oral Tablet (12.1%)  370570: amitriptyline Oral Tablet (11.8%)  373149: ondansetron Disintegrating Oral Tablet (10.7%) |  |  | 745678: albuterol Metered Dose Inhaler (16.5%)  370672: ibuprofen Oral Suspension (11.4%)  876077: calcium chloride / lactate / potassium chloride / sodium chloride Inje… (10.5%)  370509: acetaminophen Oral Suspension (10.5%) |

**Figure 5S: Heatmap of incident post-acute diagnoses by cluster, with unclustered patients shown separately, cohort B**

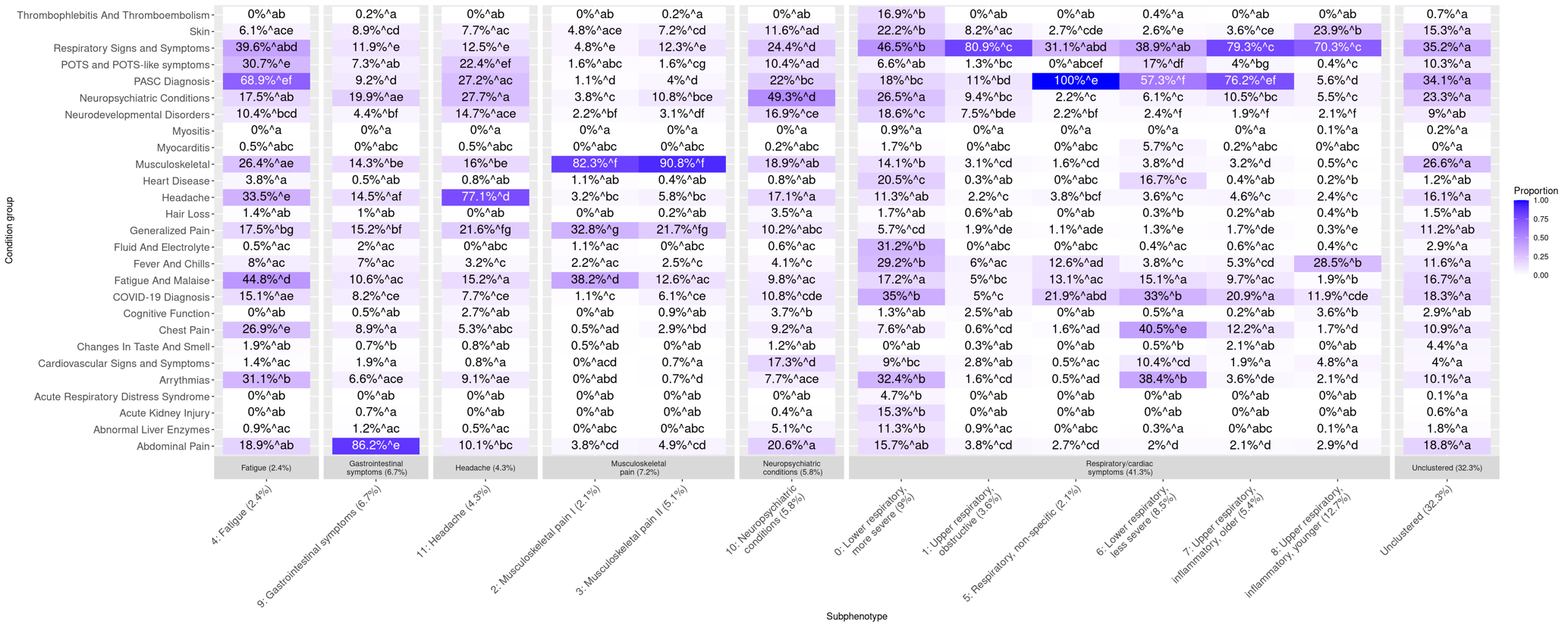

**Figure 6S: Healthcare utilization trajectories, cohort B**

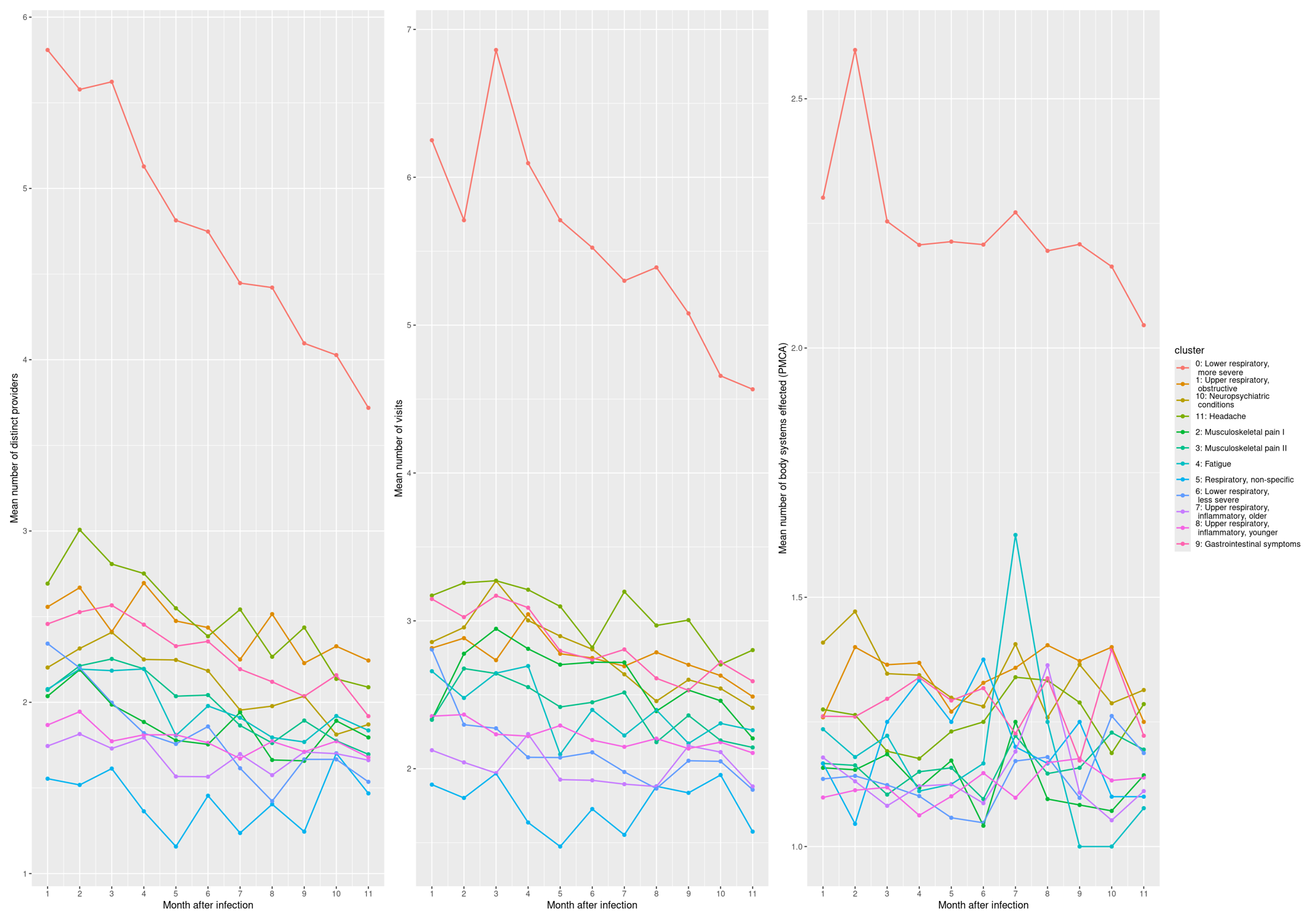

**Figure 7S: Heatmap of pre-existing chronic conditions by cluster, cohort B**

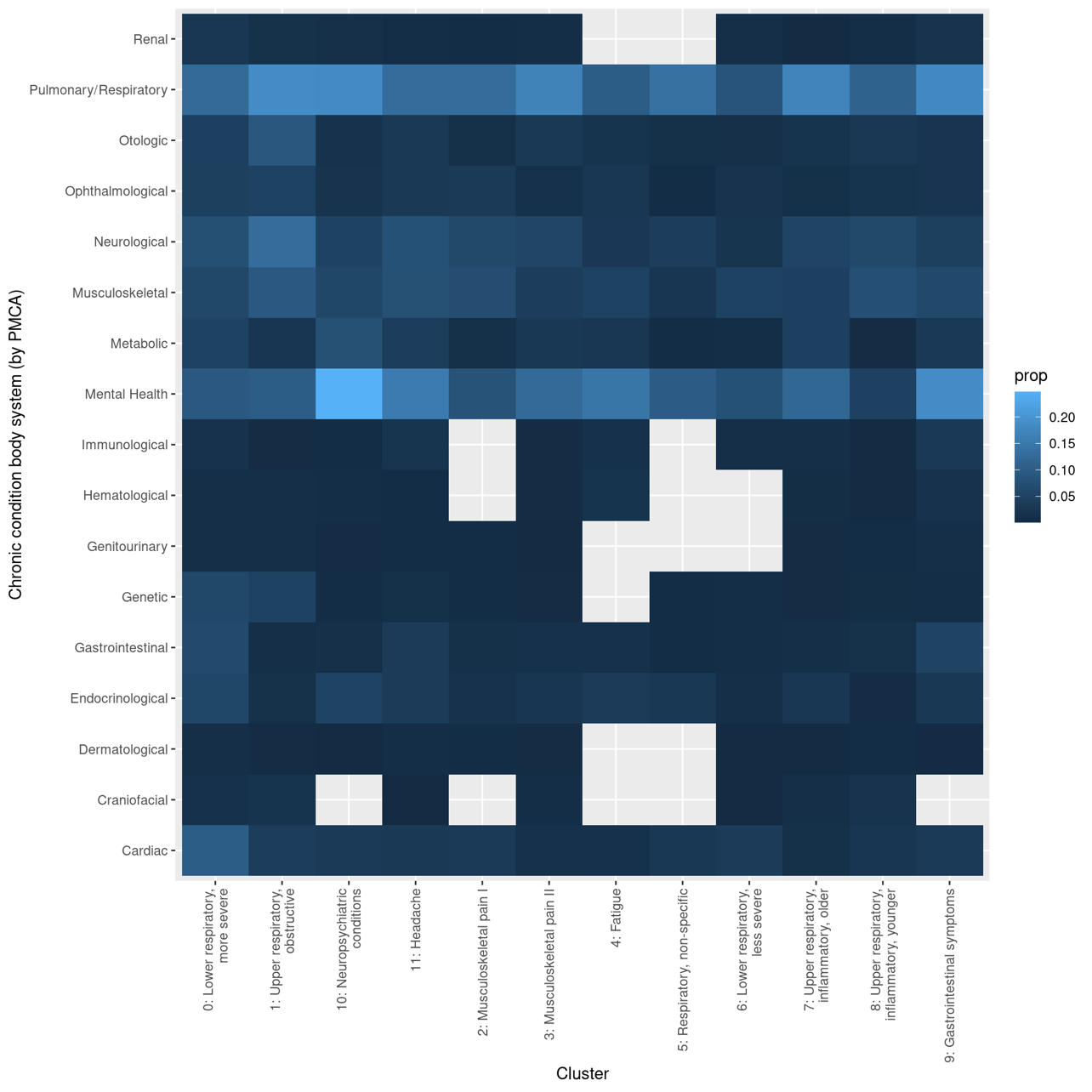
